# Antibody-dependent enhancement (ADE) of SARS-CoV-2 infection in recovered COVID-19 patients: studies based on cellular and structural biology analysis

**DOI:** 10.1101/2020.10.08.20209114

**Authors:** Fan Wu, Renhong Yan, Mei Liu, Zezhong Liu, Yingdan Wang, Die Luan, Kaiyue Wu, Zhigang Song, Tingting Sun, Yunping Ma, Yuanyuan Zhang, Qimin Wang, Xiang Li, Ping Ji, Yaning Li, Cheng Li, Yanling Wu, Tianlei Ying, Yumei Wen, Shibo Jiang, Tongyu Zhu, Lu Lu, Yongzhen Zhang, Qiang Zhou, Jinghe Huang

## Abstract

Antibody-dependent enhancement (ADE) has been reported in several virus infections including dengue fever virus, severe acute respiratory syndrome (SARS) and Middle East respiratory syndrome (MERS) coronavirus infection. To study whether ADE is involved in COVID-19 infections, *in vitro* pseudotyped SARS-CoV-2 entry into Raji cells, K562 cells, and primary B cells mediated by plasma from recovered COVID-19 patients were employed as models. The enhancement of SARS-CoV-2 entry into cells was more commonly detected in plasma from severely-affected elderly patients with high titers of SARS-CoV-2 spike protein-specific antibodies. Cellular entry was mediated via the engagement of FcγRII receptor through virus-cell membrane fusion, but not by endocytosis. Peptide array scanning analyses showed that antibodies which promote SARS-CoV-2 infection targeted the variable regions of the RBD domain. To further characterize the association between the spike-specific antibody and ADE, an RBD-specific monoclonal antibody (7F3) was isolated from a recovered patient, which potently inhibited SARS-Cov-2 infection of ACE-2 expressing cells and also mediated ADE in Raji cells. Site-directed mutagenesis the spike RBD domain reduced the neutralization activity of 7F3, but did not abolish its binding to the RBD domain. Structural analysis using cryo-electron microscopy (Cryo-EM) revealed that 7F3 binds to spike proteins at a shift-angled pattern with one “up” and two “down” RBDs, resulting in partial overlapping with the receptor binding motif (RBM), while a neutralizing monoclonal antibody that lacked ADE activity binds to spike proteins with three “up” RBDs, resulting in complete overlapping with RBM. Our results revealed that ADE mediated by SARS-CoV-2 spike-specific antibodies could result from binding to the receptor in slightly different pattern from antibodies mediating neutralizations. Studies on ADE using antibodies from recovered patients via cell biology and structural biology technology could be of use for developing novel therapeutic and preventive measures for control of COVID-19 infection.

## Introduction

The global pandemic of coronavirus disease 2019 (COVID-19), caused by severe acute respiratory syndrome coronavirus 2 (SARS-CoV-2), had resulted in a total of 34.8 million cases of infection and over 1 million deaths worldwide by October 4, 2020,^1^. No therapeutic drugs against SARS-CoV-2 are currently available, and the development of vaccines is considered as the most effective approach to control the ongoing pandemic. Multiple platforms are being developed as a SARS-CoV-2 vaccine, including DNA- and RNA-based formulations, recombinant viral subunits, replicating viral vectors and purified inactivated viral particles, are under development, and several vaccine candidates are presently being evaluated for efficacy in phase III trials^2^.

Most vaccines incorporate SARS-CoV-2 spike (S) protein or its receptor-binding domain (RBD) as immunogens. As the primary targets for neutralizing antibodies (NAbs), the S protein and RBD are promising immunogens to induce protective NAbs in vaccine recipients^3-6^. However, preclinical experience with severe acute respiratory syndrome (SARS) and Middle East respiratory syndrome (MERS) vaccine candidates has raised safety concerns about the potential for antibody-dependent enhancement (ADE) induced by coronavirus S protein^7-11^. ADE is an enhancement of viral entry into immune cells mediated by antibody via the engagement of the Fc receptors^12,13^. This phenomenon has been documented with mosquito-borne flavivirus infections, such as dengue^14^ and Zika viruses^15^. For dengue virus, the ADE of virus infection of immune cells resulted in the enhancement of disease severity especially at the second infections with different virus strains in humans^16^. For coronaviruses, ADE has been mainly reported in animal models infected by SARS-CoV, MERS-CoV and feline coronavirus, in which exacerbated lung disease was observed when vaccinated animals were infected with viruses^7,10,17^. In both SARS-CoV and MERS-CoV infections, ADE was mediated with antibodies against spike (S) proteins^7,9^. Although S protein-specific antibodies were elicited in most patients with COVID-19, the antibody titers were higher in elderly patients of COVID-19, and stronger antibody response was associated with delayed viral clearance and increased disease severity in patients^18,19^. Hence it is reasonable to speculate that S protein-specific antibodies may contribute to disease severity during SARS-CoV-2 infection^11,20,21^. Furthermore, the potential for such ADE responses is of concern for SARS-CoV-2 in the use of convalescent plasma or antibodies as a treatment in COVID-19 patients ^22,23^. However, whether SARS-CoV-2 specific antibodies or convalescent plasma could promote virus infection of immune cells or enhance disease severity has not been documented.

Here, we used an *in vitro* pseudotyped SARS-CoV-2 infection assay to evaluate the ability of plasma and antibodies from recovered COVID-19 patients to promote SARS-CoV-2 infection of immune cells, and analyzed the associated clinical and immunological characteristics.

## Results

### Clinical Characteristics

This study enrolled 222 patients in total who had recovered from COVID-19 and were discharged from the Shanghai Public Health Clinical Center as of April 23, 2020. Of the 222 patients, 205 had mild symptoms and 17 had severe symptoms. The median [interquartile range, IQR] age of patients was 53 [38-65] years; 49 % of the patients were female. The median length of hospital stay was 17 [13-24] days, and the median disease duration was 23 [18-30] days.

### Plasma from recovered patients of COVID-19 showed enhancement of SARS-CoV-2 infection of immune cells

We collected plasma samples from 205 patients who had recovered from mild COVID-19 at the time of discharge (median days 22), as well as 17 patients who had recovered from severe COVID-19 at the time before discharge (median days 34), and evaluated the enhancement of pseudotyped SARS-CoV-2 infection *in vitro* for each patient plasma by using Raji cells that are lymphoma cells derived from human B lymphocytes. The cells expressed human FcγRII (CD32) and were used for ADE assay of SARS-CoV previously^24^. Plasma from 16 (8%) of the recovered patients with mild COVID-19 and 13 (76%) of the recovered patients with severe COVID-19 (N=17, median age 66) showed a concentration-dependent enhancement of SARS-CoV-2 infection of Raji cells, indicated by the increase of luciferase expression in Raji cells (Figure S1,1A). The enhancement of virus infection was significantly higher in plasma from COVID-19 patients compared with plasma from uninfected controls (P < 0.0001, Figure 1B, S1). Moreover, plasma from these 29 patients also showed detectable enhanced infection of Raji cells of pseudotyped bat-origin SARS-like coronavirus, either RS3367 or WIV1 (P = 0.0108 or P = 0.0046, Figure 1B), while none of the plasma showed enhancement of SARS-CoV infection (Figure 1B).

**Figure 1.**
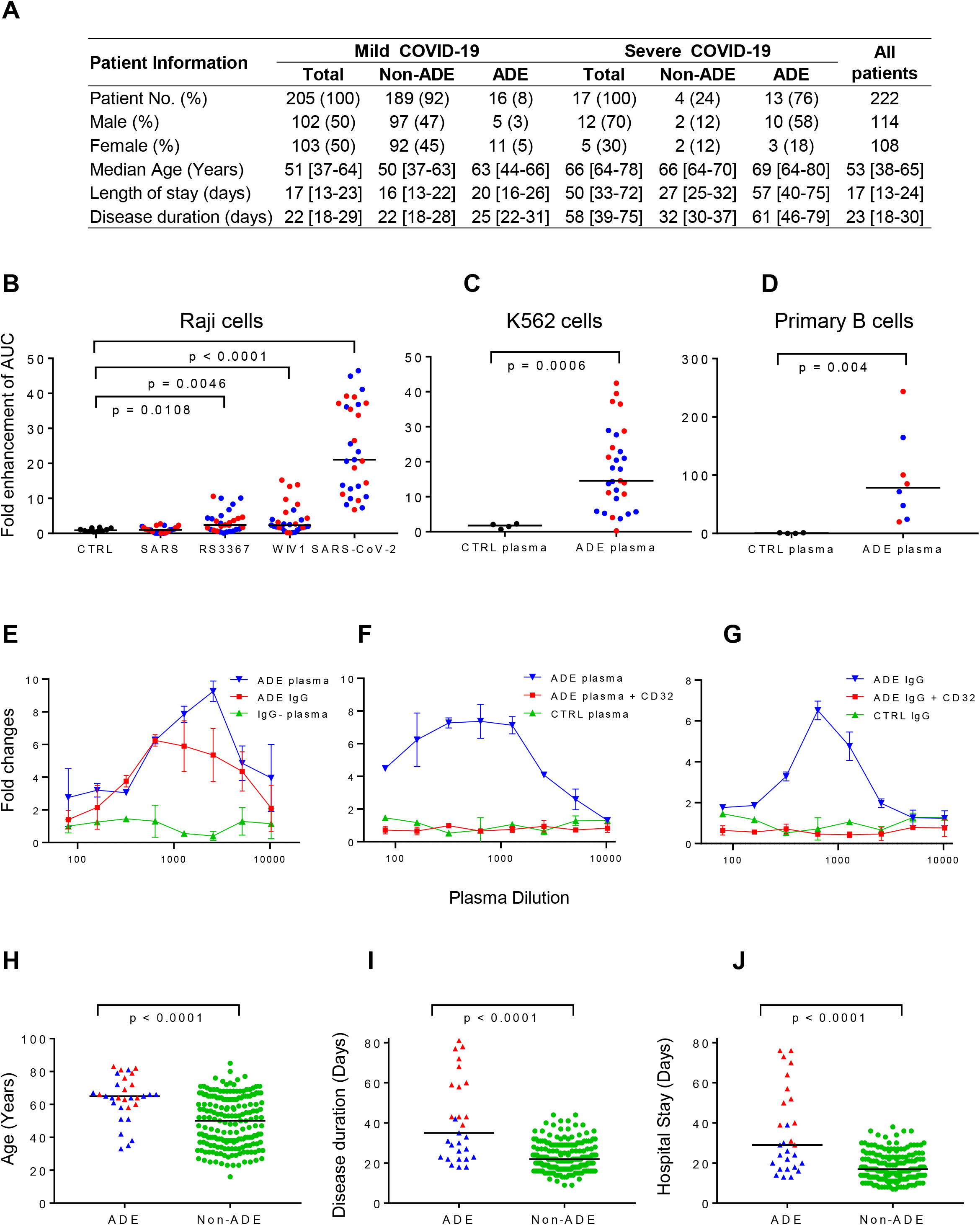
Plasma from 8% of the recovered patients with mild COVID-19 and 76% of the recovered patients with severe COVID-19 showed enhancement of SARS-CoV-2 infection through IgG Fc with the engagement of FcrRII receptor. (A) Clinical characteristics of COVID-19 recovered patients whose plasma showed ADE. (B) The enhancement of SARS-CoV-2, SARS-CoV, SARS-related RS3367, WIV-1 infection of Raji cells by 29 plasma samples from patients who recovered from mild COVID-19 (N=16, blue) or severe COVID-19 (N=13, red) are shown. Plasma from 10 uninfected donors was used as negative controls. For each plasma sample, the area under curve (AUC) of fold changes of enhancement was calculated. (C) The enhancement of SARS-CoV-2 infection of K562 cells by 29 plasma samples are shown. (D) The enhancement of SARS-CoV-2 infection of primary B cells by eight representative plasma samples are shown. (E) ADE of SARS-CoV-2 infection was mediated by plasma IgG with the engagement of FcrRII receptor. IgG purified from the ADE plasma was evaluated for enhancement of SARS-CoV-2 infection of Raji cells. IgG-depleted plasma was also evaluated. (F) ADE plasma or (G) IgG purified from ADE plasma (right) were evaluated for ADE on Raji cells in the presence or absence of anti-FcrRII antibody CD32. Comparison of age (H), disease duration (I), and hospital stay (J) of 29 patients whose plasma showed ADE effect and 193 patients whose plasma did not show ADE effect (green). P value was calculated using non-parametric *t* test.

The enhancement of SARS-CoV-2 infection by patient plasma was also observed when K562 cells derived from human monocytes were used as targets (P = 0.0006, Figure S2, 1C). Furthermore, the enhancement of SARS-CoV-2 infection was also confirmed when cultured primary B cells were used as targets. As shown in Figure S3, eight representative positive plasma samples, four from patients with mild COVID-19 and the other four from patients with severe COVID-19, showed concentration-dependent enhancement of SARS-CoV-2 infection of primary B cells. These eight plasma mediated significantly higher SARS-CoV-2 infection than control plasma from uninfected donors (P = 0.004, Figure 1D).

In the following studies, Raji cells were used as targets on the mechanism of enhancement of SARS-CoV2 infection because they were easily maintained and generated higher luciferase reading than K562 cells and primary B cells.

### Enhancement of SARS-CoV-2 infection was mediated by IgG antibodies engagement of FcγRII receptor

To confirm whether the enhancement of SARS-CoV-2 infection was mediated by antibodies, we purified IgG from the plasma and measured the enhancement of SARS-CoV-2 infection of Raji cells by purified antibodies and IgG-depleted plasma, respectively. As shown in Figure 1E, purified IgG showed enhancement of SARS-CoV-2 infection, which was similar to plasma from patients. The depletion of IgG from plasma completely abolished the infection of Raji cells, confirming that the enhancement of SARS-CoV-2 infection was mediated by IgG in plasma. We further used anti-CD32 antibody to block the cell surface FcγRII receptor to evaluate the engagement of FcγRII receptor in promoting SARS-CoV-2 infection. The addition of anti-CD32 antibody eliminated the enhancement of SARS-CoV-2 infection by both plasma (Figure 1F) and purified IgG (Figure 1G) from patients. These results indicated that the *in vitro* enhancement of SARS-CoV-2 infection by patient plasma was mediated by IgG antibodies with the engagement of FcγRII receptor, which is similar to the ADE of virus infections including SARS-CoV, MERS-CoV, Zika, and dengue viruses.

### ADE is more likely to develop in elderly patients with severe and critical condition, longer hospital stays and disease duration

We investigated the clinical characteristics of 29 recovered patients whose plasma showed ADE effect. The median age of these patients (65 [58-72] years) was significantly higher than the patients without ADE effect (50 [37-64] years, P < 0.0001, Figure 1H). The median disease duration time and the length of hospital stays of patients whose plasma showed ADE was significantly longer than patients without ADE effect (35 [23-60] days *vs*. 22 [18-29] days, P < 0.0001, and 30 [19-55] days *vs*. 17 [13-23] days, P < 0.0001, Figure 1I and 1J). These results indicated that ADE is more likely to develop in elderly patients with severe and critical condition, longer hospital stays and disease duration, suggesting a possible association of ADE with disease severity in COVID-19 patients.

To evaluate whether the ADE effect resulted from pre-exposure to other pathogens in elderly patients, we collected plasma from 18 uninfected elderly donors aged 60 to 80 years and tested them for ADE. None of the plasma from uninfected control donors showed an ADE effect (P = 0.3085, Figure S4), confirming that ADE appeared to be the result of SARS-CoV-2 infection.

### ADE is more likely to develop in patients with high titers of SARS-CoV-2 RBD-and S1-specific antibodies

Next, we evaluated the relationship between ADE effect and SARS-CoV-2-specific antibodies. Significantly higher titers of SARS-CoV-2 NAbs (P < 0.0001, Figure 2A), as well as RBD-specific (P < 0.0001) and S1-specific binding antibodies (P < 0.0001) (Figure 2B), were found in plasma with ADE effect compared to plasma without ADE effect, while S2-specific antibodies showed no difference. Then we evaluated the kinetics of ADE effect, binding antibodies, and NAbs during the course of disease in six patients for whom sequential plasma samples were available. The kinetics of ADE development was similar among all patients, starting to increase at day 10 post-disease onset, reaching their peak at day 20, and remaining stable for at least 40-81 days (Figure 2C and Figure S5A). The kinetics of titers of antibodies binding to RBD and S1 (Figure 2D and Figure S5B) was similar to the kinetics of ADE (Figure 2C and Figure S5B red line), while the kinetics of NAbs in these patients was different. The titers of NAbs in the six patients increased on day 10 post-disease onset and reached a very high level around day 20 (median ID50 = 2877) (Figure 2E and Figure S3C blue line). However, NAb titers dramatically dropped to low levels (median ID50 = 545) after day 30 post-disease onset. These results indicated that high levels of binding antibodies might contribute to the ADE of SARS-CoV-2 infection.

**Figure 2.**
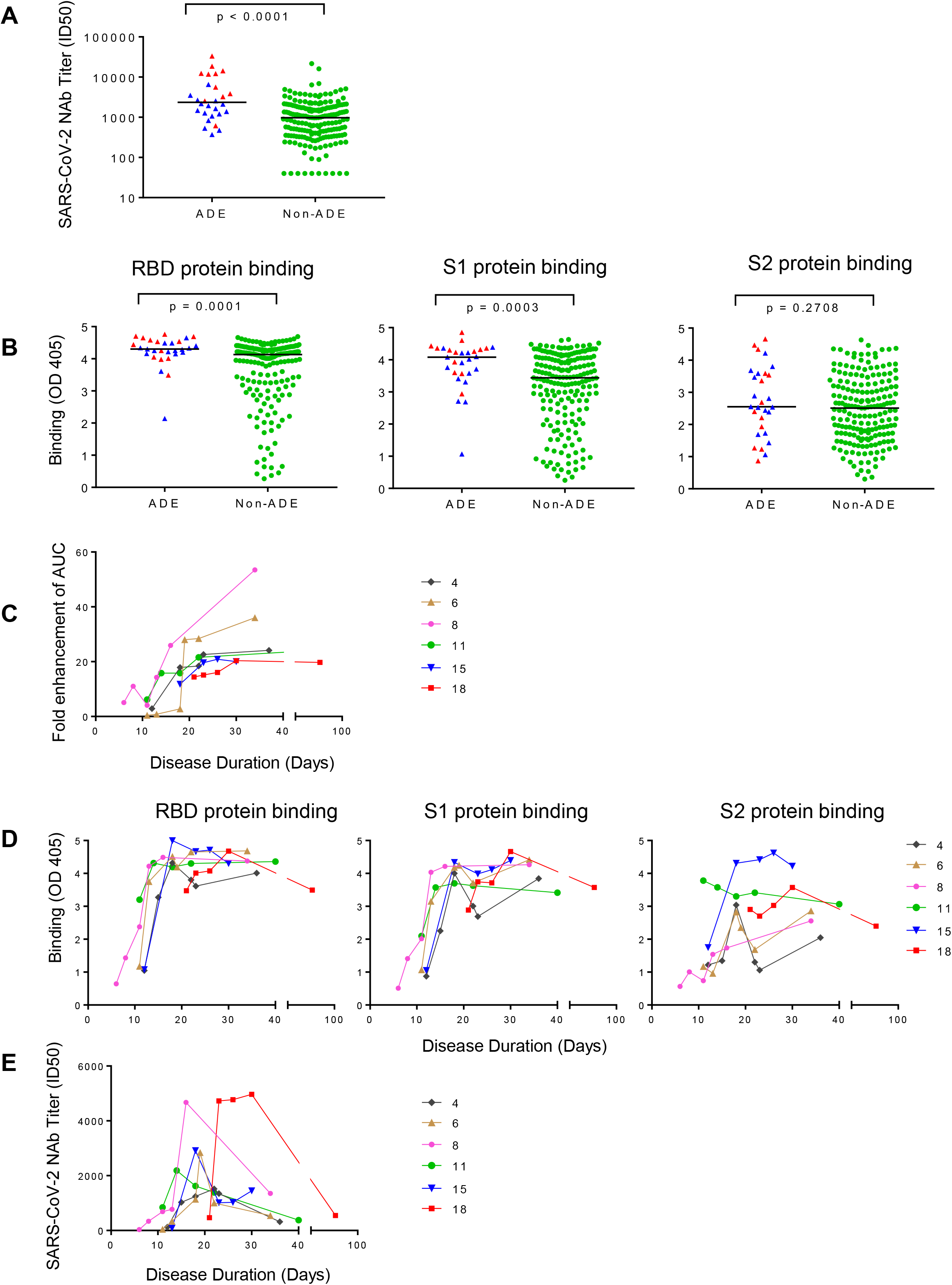
ADE is more likely to develop in elderly patients with high titers of SARS-CoV-2 RBD-and S1-specific antibodies. (A) SARS-CoV-2 NAb titers (ID50) and (B) RBD, S1-, and S2-specific binding antibodies of 29 ADE patients and 193 Non-ADE patients are compared. P value was calculated using *t* test. (C) Kinetics of SARS-CoV-2-specific ADE in plasma of six COVID-19 patients are shown. Plasma was collected at different time points post-disease onset. (D) Kinetics of spike-binding antibodies (left Y axis) targeting RBD, S1, and S2 in plasma of six COVID-19 patients exhibiting ADE are shown. Plasma diluted 1:400 was incubated with RBD, S1, or S2 protein. (E) Kinetics of SARS-CoV-2 NAbs titers in plasma of six COVID-19 patients whose plasma showed ADE are shown.

### ADE was mediated by antibodies binding to SARS-CoV-2 spike RBD subunits

To further determine the role of spike-specific antibodies in mediating ADE of SARS-CoV-2 infection, we incubated SARS-CoV-2 RBD and S1 proteins with plasma to block the protein-specific antibodies before measuring the ADE effect of plasma samples. Pre-incubation with SARS-CoV-2 RBD protein at a concentration as low as 0.1 μg/ml could completely block the ADE effect of plasma from the representative patient 8 (Figure 3A), and pre-incubation with S1 protein at the concentration of 1 μg/ml could also block the ADE effect (Figure 3B). However, pre-incubation with SARS-CoV RBD or S1 protein did not change the ADE activity in plasma (Figure 3C and 3D). The inhibition of ADE effect by SARS-CoV-2 RBD protein was also observed in the plasma from other patients. As shown in Figure 3E, pre-incubation of 10 μg/ml SARS-CoV-2 RBD significantly reduced ADE effect mediated by plasma from six tested patients (P = 0.009). These results indicated that the ADE of SARS-CoV-2 infection was mediated by antibodies targeting SARS-CoV-2 spike RBD subunits.

**Figure 3.**
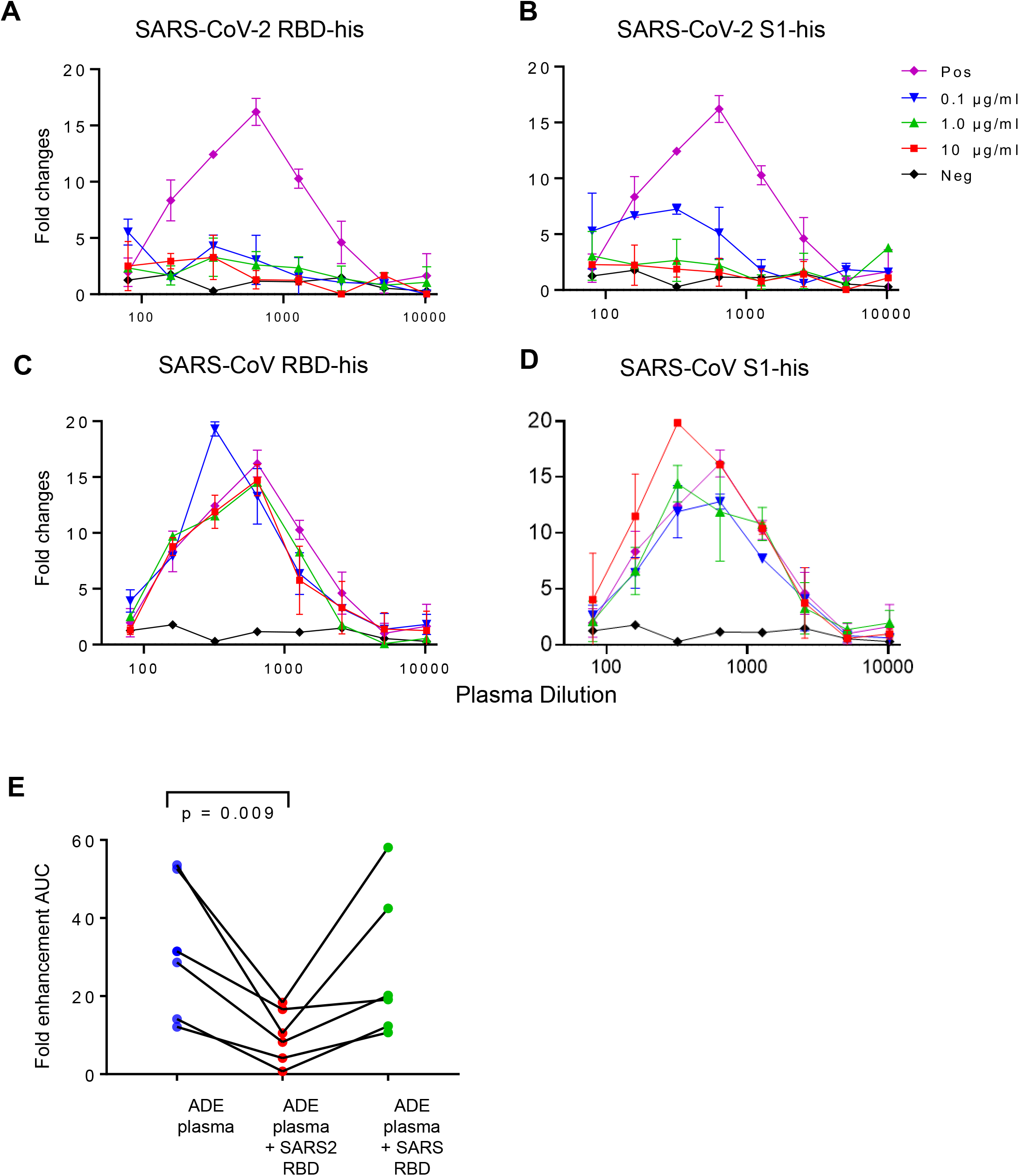
ADE was mediated by antibodies binding to SARS-CoV-2 spike RBD subunits. (A) Suppression of ADE of plasma by RBD (A) or S1 protein (B) of SARS-CoV-2 but not by RBD (C) or S1 protein (D) of SARS-CoV viruses. Serially diluted patient plasma were pre-incubated with different concentrations of proteins before evaluating ADE of SARS-CoV-2 infection of Raji cells. Untreated plasma was used as a positive control, and healthy donor plasma was used as a negative control. (E) ADE mediated by plasma from six patients was inhibited by pre-incubation with 10 μg/ml SARS-CoV-2 RBD but not SARS-CoV RBD.

### ADE of SARS-CoV-2 infection occurred through the virus-cell membrane fusion

It was suggested that the ADE of viral infection was mediated by phagocytosis of immune complexes via FcγRII / CD32 receptor^25^. However, the addition of chloroquine, a phagocytosis inhibitor which could raise the pH of phagolysosomes and inhibit the phagocytosis of mononuclear cells^26^, did not inhibit the ADE by the plasma, even at the highest concentration of 50 μM (Figure 4A). In contrast, EK1 peptide, which has been demonstrated to inhibit virus-cell membranes fusion by binding to the HR1 domain and thus inhibiting the formation of the six-helix bundle (6HB) of SARS-CoV-2 S2 protein^27^, blocked ADE of SARS-CoV-2 infection in a dose-dependent manner (Figure 4B). The inhibition of ADE effect by EK1 peptide, but not chloroquine, was observed for six tested plasma samples with ADE effect (P = 0.0064, Figure 4C). These results indicated that ADE of SARS-CoV-2 infection was mediated through virus-to-cell membrane fusion, not phagocytosis.

**Figure 4.**
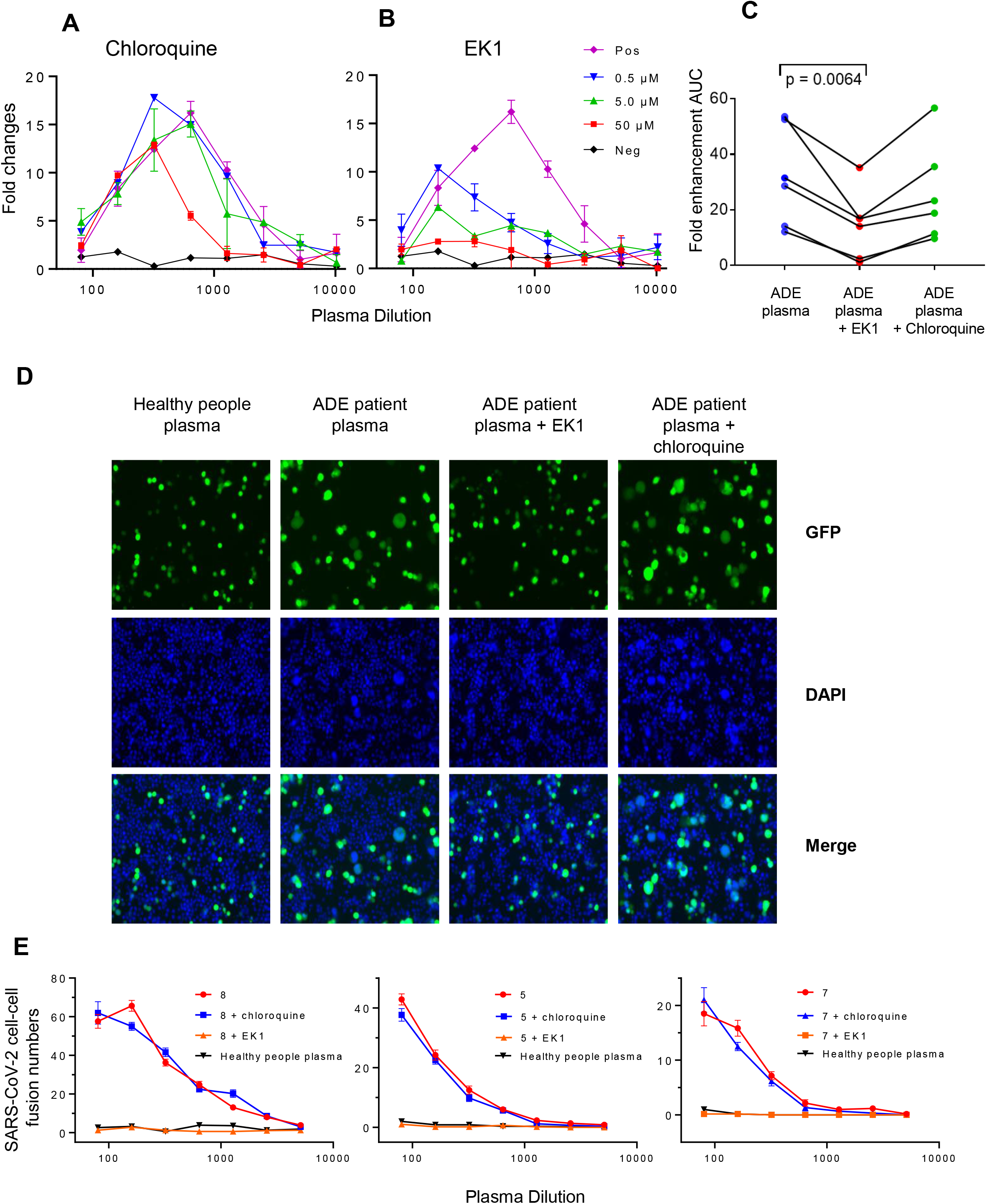
ADE-mediated SARS-CoV-2 entry into cells is through virus-cell membrane fusion. Inhibition of ADE induced by plasma from patients using chloroquine (A) or EK peptide (B). Serially diluted plasma was pre-incubated with different concentrations of chloroquine or EK1 peptide before evaluating ADE of SARS-CoV-2 infection of Raji cells. Patient plasma with ADE was used as a positive control, and plasma from uninfected health donor was used as a negative control. (C) ADE mediated by plasma from six patients was inhibited by 50 μM of EK1 peptide but not by chloroquine.. (D) ADE plasma from recovered COVID-19 patients promoted syncytium formation. Plasma from uninfected health donor was used as a negative control. The syncytium formation was specifically inhibited by EK1 peptide but not by chloroquine. (E) The statistical counts of syncytium formation induced by plasma from three patients (8, 5, and 7) in the presence of EK1 peptide or chloroquine.

The ability of plasma to promote virus-to-cell membrane fusion was confirmed by an *in vitro* syncytium formation assay, using HEK-293T cells expressing the SARS-CoV-2 S protein as effector cells and Raji cells as target cells. No syncytium formation occurred in the presence of control plasma (Figure 4D, left). However, large syncytium was induced by plasma from representative patients 5, 7, and 8 in a dose-dependent manner (Figure 4D, middle; Figure 4E). The syncytium formation induced by plasma from COVID-19 patients was specifically inhibited by the addition of EK1 peptide, but not chloroquine (Figure 4D, right; Figure 4E). These results again confirmed that ADE of SARS-CoV-2 infection by plasma was mainly through cell-to-cell membrane fusion, a pathway involved in the formation of the six-helix bundle (6HB) of SARS-CoV-2 S2 protein, which could be inhibited by EK1 peptide.

### RBD-specific human NAb 7F3 with ADE effect enhanced SARS-CoV-2 infection and promoted virus entry into Raji cells through the virus-cell membrane fusion

We further evaluated the characterization of antibodies with ADE effect by isolating two monoclonal antibodies (mAbs) from recovered COVID-19 patient. These two mAbs, termed as 7F3 and 4L12, were isolated by *in vitro* single B cell culture and subsequent high-throughput micro-neutralization screening assay from the same recovered COVID-19 patient. Both of the antibodies potently neutralized SARS-CoV-2 pesudovirus infection of 293T cells expressing ACE2 protein with an IC50 of 0.00684 μg/ml and 0.00452 μg/ml, respectively (Figure 5A). The two antibodies bound to SARS-CoV-2 RBD and S1 proteins, but not S2, in the ELISA assay (Figure 5B). Antibody 7F3 had higher binding affinity to RBD protein with a KD value of 0.69± 0.03 nM, when compared to antibody 4L12 which bound to RBD with a KD value of 1.49 ± 0.06 nM (Figure 5C). Antibody 7F3 showed a concentration-dependent enhancement of SARS-CoV-2 infection of Raji cells (Figure 5D), while antibody 4L12 did not. The enhancement of SARS-CoV-2 infection by antibody 7F3 was also dependent on the interaction between antibody Fc region with FcγRII receptor, because the enhancement could be completely abolished by either removal of antibody Fc region (Figure 5E) or blocking FcγRII receptor with anti-CD32 antibody (Figure 5F). We also compared the ADE effect of different isotypes of 7F3 antibodies which were generated by linking different heavy-chain constant regions to the same variable region of 7F3 antibody. The IgG1 isotype showed the strongest ADE effect, IgG4 isotype showed detectable ADE effect, while IgG2 and IgG3 did not show any detectable ADE (Figure 5G), possibly resulting from the different binding affinity to FcγRII receptor on Raji cells. Consistent with the observations for plasma samples from recovered patients, antibody 7F3-mediated enhancement of SARS-CoV-2 infection could be specifically reduced by pre-incubation with RBD protein of SARS-CoV-2, but not from SARS-CoV virus (Figure 5H), and the ADE could also be blocked by fusion-inhibitor EK1 peptide, but not chloroquine (Figure 5I), suggesting that antibody 7F3-mediated ADE of SARS-CoV-2 infection of Raji cells also occurred through virus-to-cell membrane fusion.

**Figure 5.**
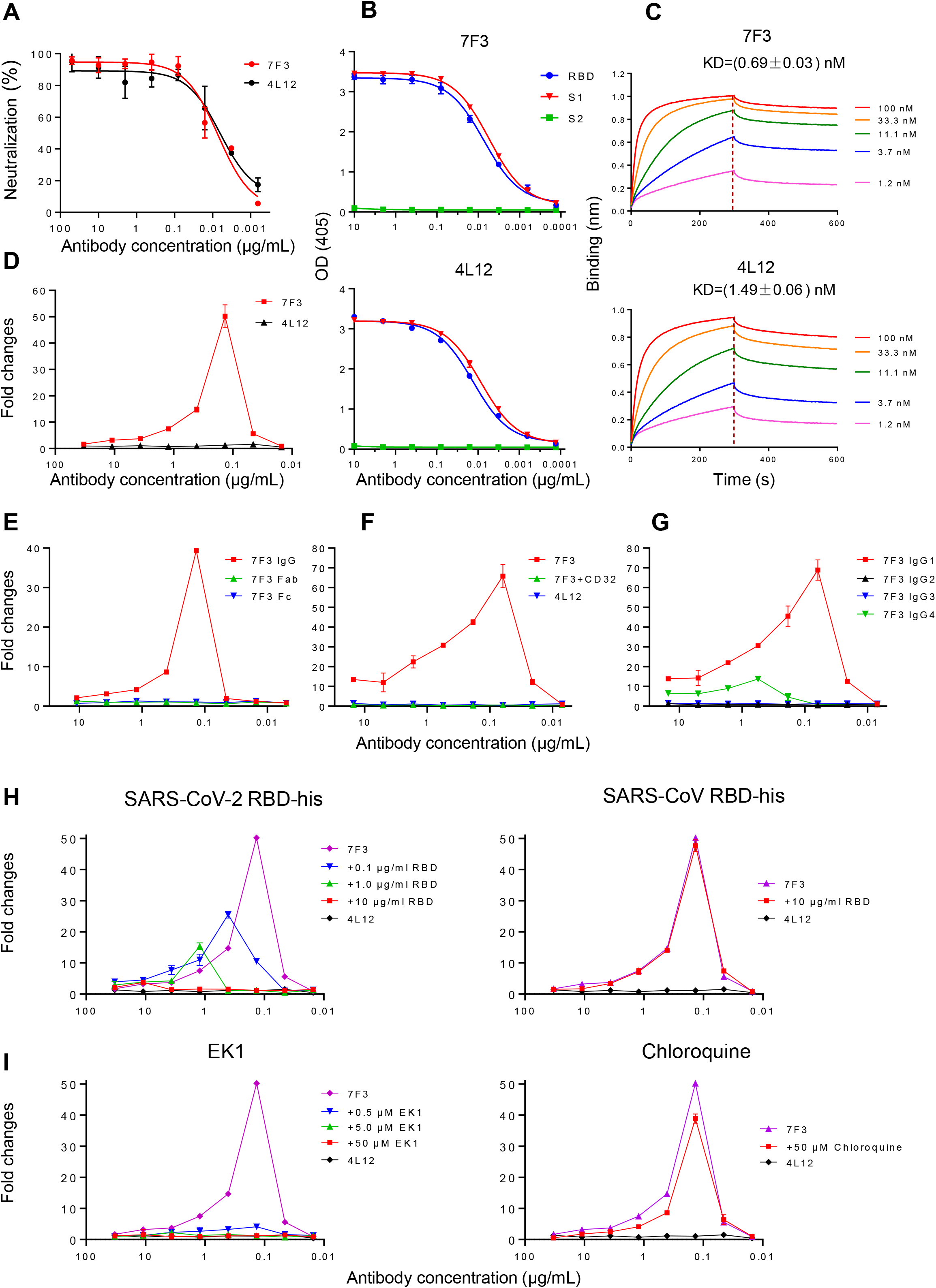
RBD-specific NAb 7F3 from a recovered patient of COVID-19 enhanced SARS-CoV-2 entry into cells through virus-cell membrane fusion. (A) Neutralizing curve of 7F3 and 4L12 against SARS-CoV-2 pseudovirus. (B) Binding of 7F3 and 4L12 mAb to SARS-CoV-2 RBD, S1, and S2 proteins in ELISA assay. (C) Binding affinity of 7F3 and 4L12 to RBD was measured by bilayer interferometry experiments. (D) Enhancement of SARS-CoV-2 infection of Raji cells by 7F3 but not by 4L12. (E) Impact of IgG, Fab, and Fc of 7F3 on ADE of SARS-CoV-2 infection. (F) Impact of IgG1, IgG2, IgG3, and IgG4 isotypes of 7F3 on ADE of SARS-CoV-2 infection. (G) Blockage of RcrRII with anti-CD32 antibody (5 μg/mL) inhibited ADE induced by 7F3. (H) RBD of SARS-CoV-2 (left) blocked ADE induced by 7F3. RBD of SARS-CoV (right) was used as a negative control. (I) Blockage of ADE induced by 7F3 with EK1 (left) and chloroquine (right).

### Peptide scanning for hot spots in RBD associated with ADE effect

Next, we explored the epitopes in RBD to which patient plasma and NAb 7F3 bound and induced ADE. We synthesized a series of 20-mer peptides with 10 amino acid overlap spanning the RBD region (304-593) to block the ADE of patient plasma and antibody 7F3. Peptides from the S1 region, i.e., 304-323, 364-383, 544-563, 564-583, and 574-593, dramatically blocked the ADE of both patient plasma (Figure 6A) and 7F3 (Figure 6B) and decreased >70 % of AUC (Figure 6C). Peptides from S1 regions 454-473 and 484-503 decreased 50-62% of the AUC for both ADE patient plasma and 7F3. Peptides from S1 regions 484-403 and 525-543 specifically blocked ADE patient plasma, but not 7F3. These results suggested that several epitopes in RBD were associated with ADE by antibodies.

**Figure 6.**
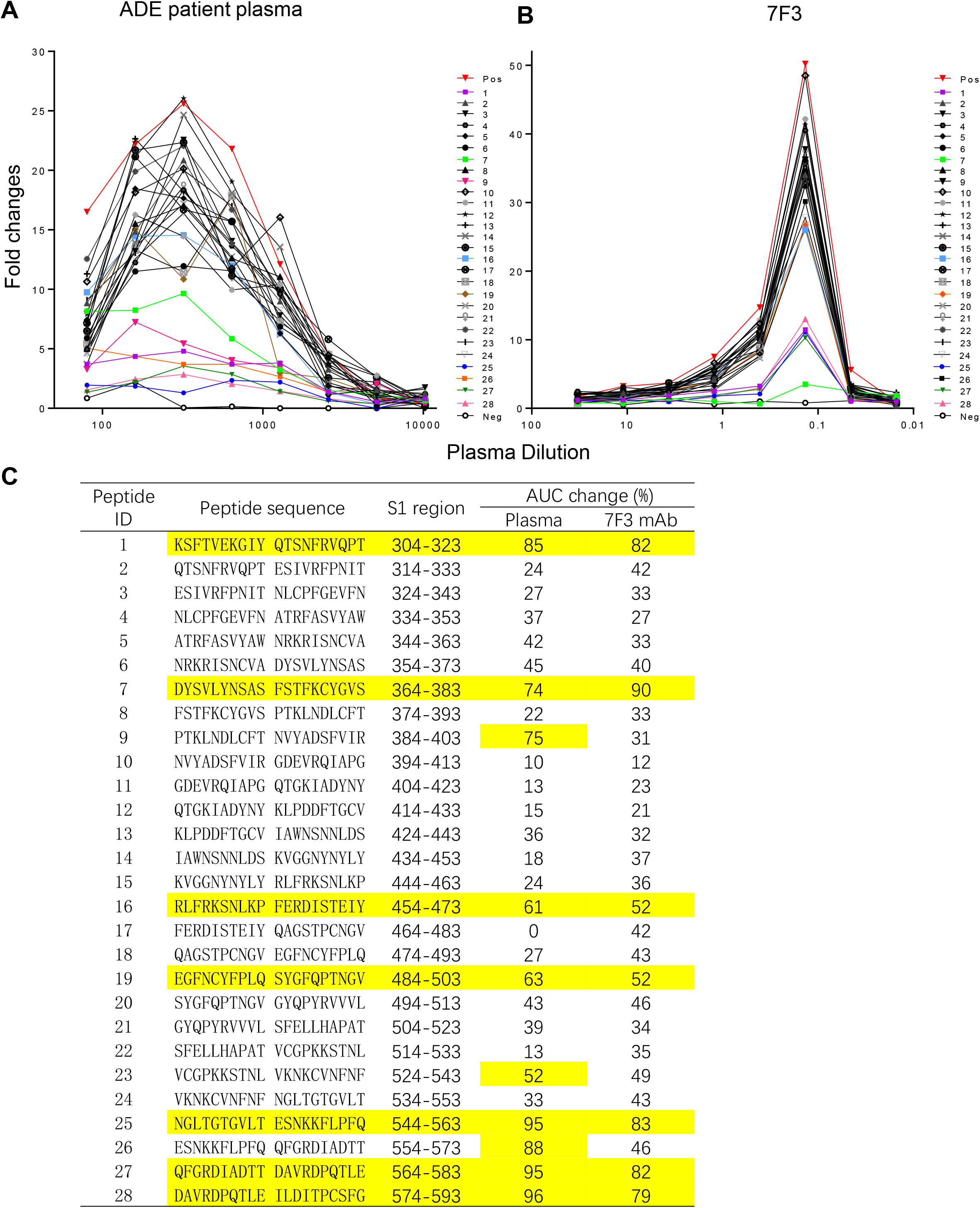
Peptide scanning for hot spots in RBD associated with ADE effect. ADE of patient plasma (A) and antibody 7F3 (B) were blocked with 20-mer overlapping peptides covering RBD protein. Serially diluted ADE patient plasma was pre-incubated with 20 μg/ml peptides before evaluating ADE effect on Raji cells. Patient plasma with ADE was used as a positive control, and plasma from uninfected health donor was used as a negative control. (C) Percentage of AUC changes of ADE curves after blocking of ADE patient plasma and 7F3 with 20-mer RBD overlapping peptides.

### ADE antibody 7F3 and non-ADE antibody 4L12 shared overlapping epitopes but showed different binding abilities to RBD

To more precisely map the epitopes on RBD recognized by antibody 7F3, we introduced single amino acid substitutions into the spike RBD domain and constructed 25 spike mutants, including seven mutants that were reported to be resistant to NAbs^28^, as well as a prevalent mutant D614G^29^ (Figure 7, highlighted in blue), and evaluated their sensitivity to neutralization of antibody 7F3. As shown in Figure 7A, 7F3 neutralized all seven mutants that resistant to NAbs and the prevalent mutant D614D. Three amino acid substitutions, including F342L, P491A, and E516A exhibited complete resistance to the neutralization of both 7F3 and 4L12 (IC50 >50 μg/ml), suggesting overlapping between the epitopes of 7F3 and 4L12.

**Figure 7.**
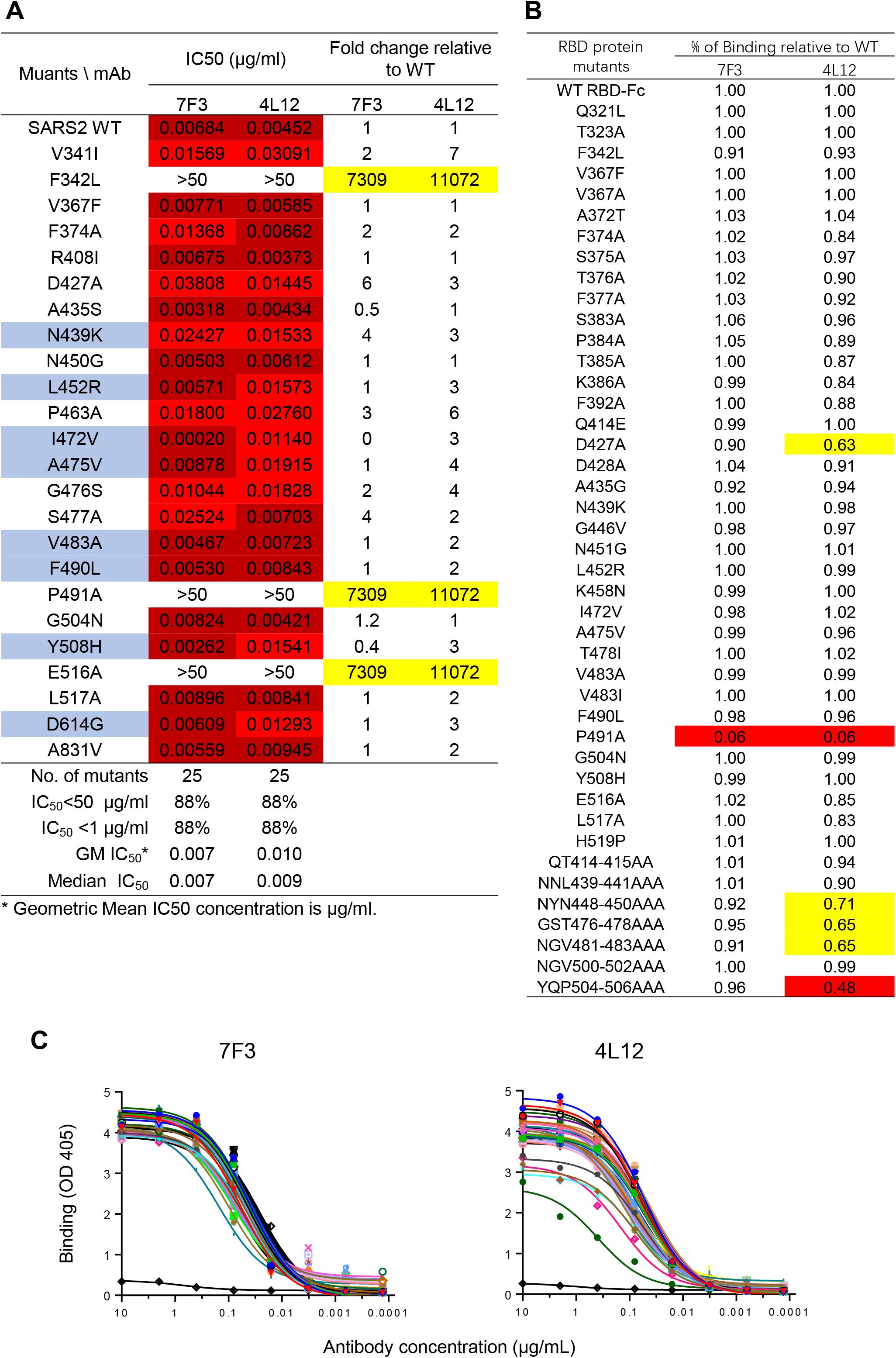
ADE antibody 7F3 and non-ADE antibody 4L12 shared overlapping epitopes but showed different binding abilities to RBD. (A) Neutralization potency and breadth of antibodies against 27 SARS-CoV-2 RBD mutants. Mutants reported to be resistant to SARS-CoV-2 NAbs were highlighted in blue. (B) Percentage of 7F3 or 4L12 binding to SARS-CoV-2 RBD mutants compared with RBD WT. Values between 0.5 - 0.8 are highlighted in yellow, and values < 0.5 are highlighted in red. (C) Binding curves of 7F3 or 4L12 to SARS-CoV-2 RBD and its mutants.

We expressed the RBD protein mutants and measured the binding ability of 7F3 to these mutants relative to wild type RBD. None of these mutations except P491A affected 7F3 and 4L12 binding to spike protein (Figure 7B and 7C). A mutant with single mutation D427A and four mutants with three amino acid alanine substitutions in RBD showed decreased binding to 4L12 but had no effect on 7F3 binding (Figure 7B and 7C). Even though mutations F342L and E516A in the RBD region affected 7F3 neutralization, they did not impact 7F3 binding, which may play an important role in ADE.

### Structures of 7F3 or 4L12 in complex with the S protein of SARS-CoV-2 revealed different binding patterns

To characterize the molecular details of the antibodies mediating ADE, we solved the cryo-EM structures of S-ECD bound with 7F3 or 4L12 at an overall resolution of 3.3 Å and 3.0 Å, respectively (Figure S6-S8, Table S1). Details of cryo-EM sample preparation, data collection and processing, as well as model building, can be found in the Materials and Methods section in Supplementary Information (SI).

The overall resolution for S-ECD was good enough for model building, whereas the resolution at the interface between 7F3 and S-ECD was worse owing to the flexibility. We only docked the light chain and heavy chain of 7F3 into the cryo EM map. The S protein bound with 7F3 exhibits a conformation with one “up” and two “down” RBDs, among which the “up” RBD and one of two “down” RBDs were bound by 7F3, whereas the other “down” RBD was not bound by 7F3 (Figure 8A). In contrast to the S/7F3 complex, all three RBDs of S protein were in “up” conformation and bound with 4L12 in the S/4L12 complex (Figure 8B). Additionally, the interfaces between antibodies and RBD in both antibodies are overlapped with binding to ACE2 (Figure S9).

**Figure 8.**
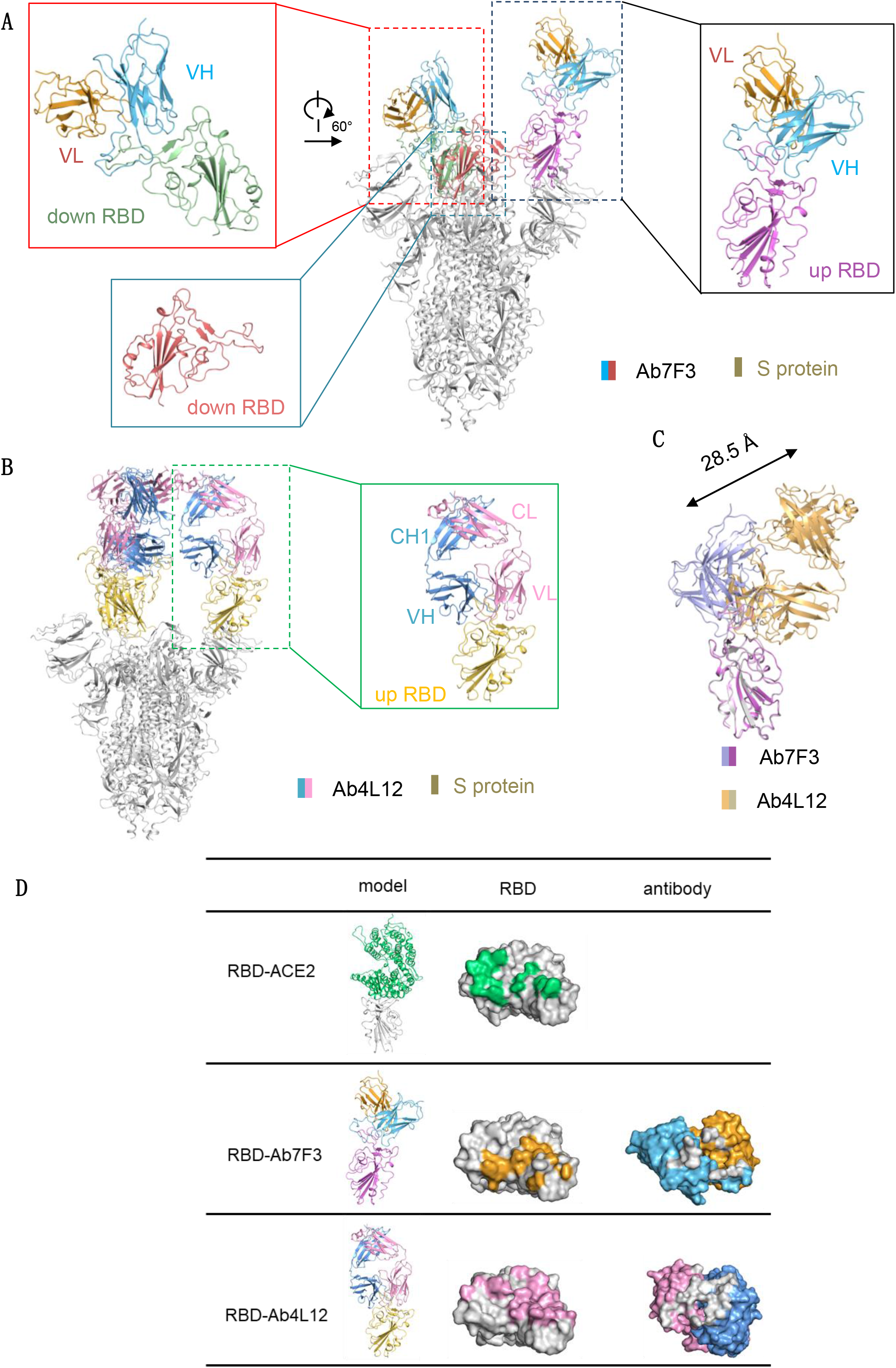
Structures of 7F3 or 4L12 in complex with the S protein of SARS-CoV-2 revealed different binding patterns. (A) Cryo-EM structure of the 7F3-bound S protein. 7F3 binds to one “up” RBD and one “down” RBD. (B) Cryo-EM structure of the 4L12-bound S protein. The insets in (A) and (B) show the details of the corresponding regions. (C) 7F3 shifts for about 28.5 angstroms compared with 4L12. (D) The ACE2 binding site and the epitopes of 7F3 and 4L12 on RBD of the S protein. The complexes of RBD with ACE2 and the antibody 7F3 and 4L12 are shown as cartoon in left column. The RBD is shown as grey surface in middle column, with the ACE2 binding site colored in green and the epitope of 7F3 and 4L12 colored in orange and pink, respectively. In the right column, the antibodies are shown as surface, with the heave and light chains colored as in (A) and the paratopes colored in grey.

The resolution at the interface between 4L12 and RBD was improved to 3.5 Å by focused refinement, allowing detailed analysis (Fig. S7). When compared with 4L12 bound structure, 7F3 bound to RBD with a shift of about 28.5 angstroms (Figure 8C), making a different binding pattern. In summary, structural analysis indicates that both 7F3 and 4L12 can block the binding between ACE2 and RBD. The binding interface of 7F3 is accessible on the “down” RBD and is partially overlapped with the edge of the receptor binding motif (RBM) (Figure 8D), which is consistent with the competing results of peptides 454-473 and 484-503 (Figure 6C) and the amino acid P491A substitution result (Figure 7A, B). Additionally, the epitope residues of 4L12 are distributed across RBM, fully competing with ACE2 (Figure 8D). These results suggest that the different ability of antibody 7F3 and 4L12 to induce ADE may result from the different binding patterns to spike proteins.

## Discussion

The role of antibodies during SARS-CoV-2 infection has remained unclear. For most infectious viral diseases, the concentrations of virus-specific antibodies correlate with viral clearance and protection, while it is different in patients of COVID-19. It was reported that stronger antibody response was associated with delayed viral clearance and increased disease severity in patients of COVID-19^30^. We also reported that NAb titers were higher in elderly patients of COVID-19, who tend to have worse outcomes, while a few patients recovered without generating detectable NAbs^18^. Here we reported the observation of *in vitro* ADE of SARS-CoV-2 entry into FcγRII receptor-bearing cells by plasma and antibodies from patients who recovered from COVID-19. The antibody enhancement of SARS-CoV-2 entry may enhance viral replication in immune cells, since it has been reported that SARS-CoV-2 could productively infect immune cells including monocytes and B cells both *in vivo* and *ex vivo*^*31*^. Because of the limited availability of tissue samples from these patients, we could not directly evaluate the immuno-pathological damage associated with ADE. However, in our study the enhancement of virus infection was more commonly observed in plasma from older patients with severe symptoms, and it was associated with prolonged disease duration, suggesting that ADE may be associated with worse clinical outcomes during SARS-CoV-2 infection.

Previous studies on SARS-CoV have shown that antibodies mediating ADE of SARS-CoV infection were mainly targeting an immunodominant linear epitope (S597–603) located at C-terminal domain of SARS-CoV spike protein^32^. Here we found that antibodies mediating enhancement of SARS-CoV-2 infection were mainly targeting the RBD domain of SARS-CoV-2 spike protein. The enhancement could be completely blocked by pre-adsorption of RBD-specific antibodies in plasma with RBD protein. As the receptor binding site of the spike protein, the RBD domain is the main target for neutralizing antibodies^33^. Our results indicated that some RBD-specific antibodies, for example antibody 7F3 in this study, have dual effects in mediating both neutralization and ADE. The effect of neutralization or ADE was dependent on receptor expression on the target cells and concentration of the antibody. When viruses infect cells expressing ACE2, such as Huh-7 cells or lung alveolar epithelial cells, antibody 7F3 at optimal neutralizing concentration could block RBD binding to ACE2 and inhibit viral infection. However, when viruses infect cells expressing Fc receptors, such as Raji, K562, or primary immune cells, the antibody at suboptimal neutralizing concentration promotes virus entry into cells through interaction between antibody and Fc receptors (Figure 9). We found that amino acid substitutions F342L and E516A on RBD allowed the virus escape from the neutralization by 7F3 without reducing binding affinity to antibody. How these mutants abolished the antibody neutralization without affecting binding affinity requires further studies.

**Figure 9.**
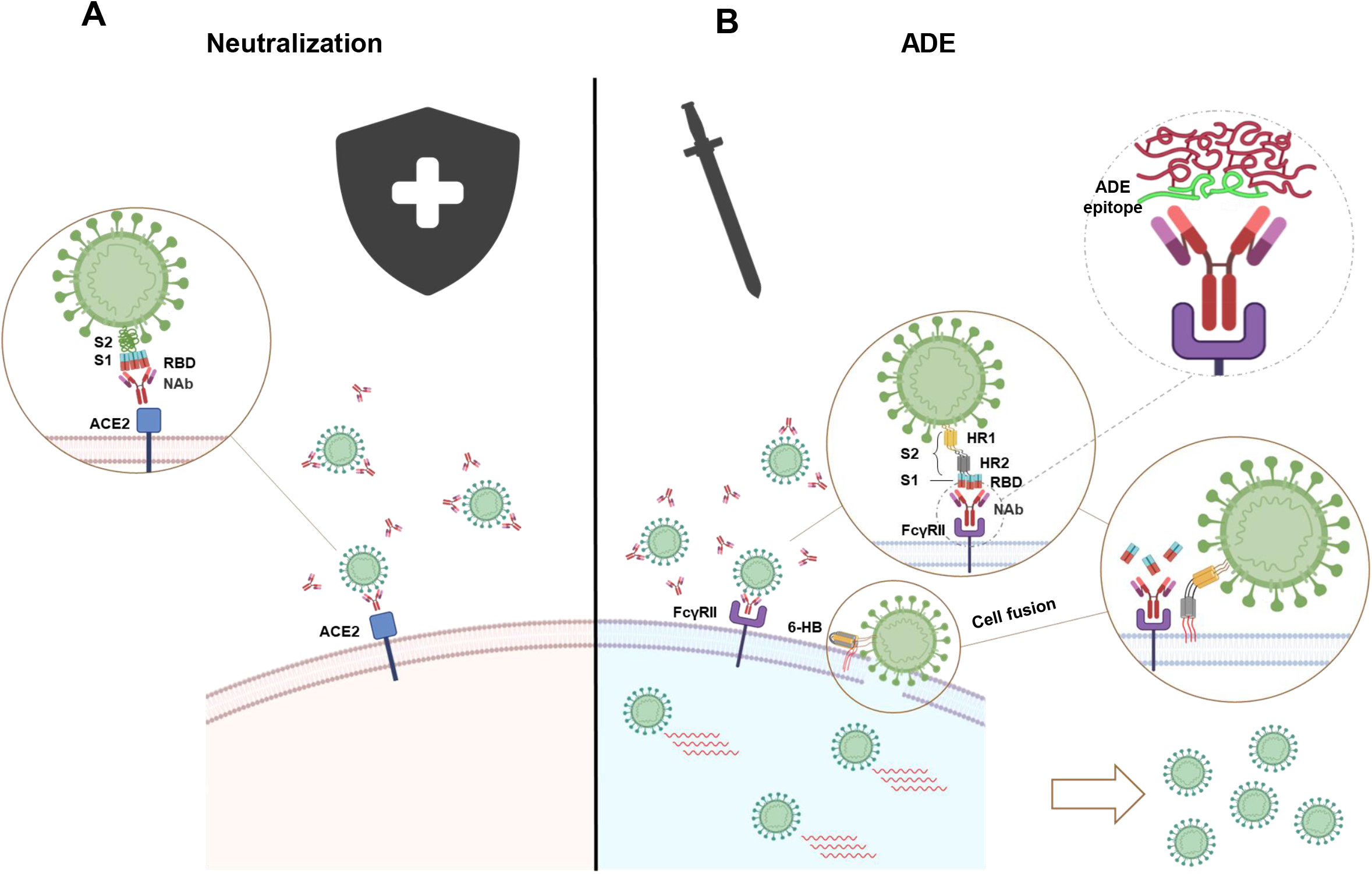
Scheme of dual effects of SARS-CoV-2 neutralizing antibodies on mediating both neutralization and ADE. (A) Binding of NAbs to RBD of SARS-CoV-2 spike protein blocks virus from infecting ACE2 receptor expressing cells. (B) Binding of NAbs to Fc receptors (FcRs) expressing immune cells with their Fc domains and to the RBD with their Fab domains triggers fusion-based viral infection.

It is interesting that antibody-mediated viral entry into Fc receptor-bearing cells was not through phagocytosis, but rather, through virus-to-cell membrane fusion. However, the molecular mechanism that regulates the interaction among spike protein, antibody and Fc receptors in order to initiate virus-cell membrane fusion remains unknown. It should be noted that not all RBD-specific antibodies will induce ADE effect. Antibodies that can induce ADE in this study bind to the spike with one “up” and two “down” RBD domains, while the antibodies that cannot induce ADE bind to the spike with three “up” RBD domains. Therefore, the different binding pattern to spike proteins may result in different abilities to promote ADE, but the detailed mechanism requires further studies.

Our results revealed that antibodies mediating ADE of SARS-CoV-2 infection were not the result of pre-existing cross-reactive antibodies from other coronavirus infection^34^, but were generated *de novo* following infection with SARS-CoV-2. First, the plasma from COVID-19 patients did not significantly promote the enhancement of SARS-CoV coronavirus infection. Second, pre-incubation with SARS-CoV RBD did not block the enhancement of virus infection by either plasma or monoclonal antibody 7F3. Third, mAb 7F3, which promotes the enhancement of virus infection, specifically binds to RBD of SARS-COV-2 virus, but no other coronaviruses. These results also suggest that ADE may be more likely to occur at later time points after recovery from COVID-19 when the concentration of neutralizing antibodies elicited by the primary SARS-CoV-2 infection have waned to suboptimal neutralizing level.

### Limitations

In this study, plasma and antibodies was measured by an *in vitro* cell-based pseudovirus assay to evaluate the enhancement of SARS-CoV-2 infection of immune cells. Whether such enhancement of virus infection results in disease severity needs to be validated in appropriate animal models.

### Implication for SARS-CoV-2 vaccine research and therapies

Although several SARS-Cov-2 vaccines have been undergoing phase III clinical trials, the potential ADE of coronavirus infection still remains a safety concern for any vaccine candidates. The observation of enhancement of SARS-CoV-2 infection mediated by plasma and antibodies from recovered COVID-19 patients in this study does not indicate that vaccine candidates would necessarily induce ADE or disease severity. However, these results suggest that vaccine candidates should be evaluated for induction of ADE in addition to induction of neutralizing antibodies. A vaccine that can induce high titers of neutralizing antibodies should be safer than one inducing low titers since 1) most the newly invaded virions are neutralized before the ADE occurs and 2) neutralizing antibodies mediate ADE only at the suboptimal neutralizing concentration. Furthermore, these results also suggested that plasma and antibodies from patients who recovered from COVID-19 should be tested for potential ADE effect before clinical usage.

## Methods

### Study design and participants

The study was conducted under a clinical protocol approved by the Investigational Review Board in the Shanghai Public Health Clinical Center (Study number: YJ-2020-S018-02). The study included a cohort of 222 adult COVID-19 recovered patients who were quarantined and hospitalized at the Shanghai Public Health Clinical Center. All patients were diagnosed with laboratory-confirmed SARS-CoV-2 infection by positive results of reverse transcriptase–polymerase chain reaction (RT-PCR) testing of nasopharyngeal samples. 205 patients were categorized as mild symptoms, and 17 patients were in severe and critical condition according to the Guidelines on the Diagnosis and Treatment of Novel Coronavirus issued by the National Health Commission, China. All participants signed an informed consent approved by the IRB. All patients had recovered and were discharged after meeting effective national treatment standards.

### Materials

The human primary embryonic kidney cell line (HEK293T) (CRL-3216™) and Raji cells were obtained from the American Type Culture Collection (ATCC). 293T cells expressing human angiotensin converting enzyme II (ACE2) (293 T/ACE2) were constructed as previously described^18^. CD19+IgA−IgD−IgM− Primary B cells were sorted out from peripheral blood mononuclear cell (PBMC) of recovered patients of COVID-19 and expanded in vitro for 13 days in the presence of irritated 3T3-msCD40L feeder cells, IL-2 and IL-21 as previously described^35^. Raji cells and K562 cells were cultured in RPMI 1640 medium with 10% fetal bovine serum (FBS), and the other cells were cultured in Dulbecco’s Modified Eagle’s Medium (DMEM) with 10% FBS. HEK293 cells expressing SARS-CoV-2 RBD protein was purchased from GenScript Company (Nanjing, China). SARS-CoV-2 S1 and S2 proteins, as well as SARS-CoV S1 and RBD proteins were purchased from Sino Biological Company (Beijing, China). The 20-mer peptides with 10 amino acid overlap spanning the entire RBD region and EK1 peptide (SLDQINVTFLDLEYEMKKLEEAIKKLEESYIDLKEL) were synthesized by Jetide (Wuhan, China). The expression plasmids for SARS S protein, pcDNA3.1-SARS-S (GenBank accession: ABD72979.1), SARS-CoV-2 S protein, pcDNA3.1-SARS-CoV-2-S (GenBank accession: NC_045512), and pcDNA3.1-RS3367 (GenBank accession: KC881006) were synthesized by Genscript. The HIV-1 Env-deficient luciferase reporter vector pNL4-3. Luc. R-E-and 3T3mCD40L cells were obtained through the NIH AIDS Reagent Program. Chloroquine was purchased from TargetMol. Pseudoviruses of SARS-CoV-2, SARS-CoV, Bat-SL-RS3367 and WIV1 coronaviruses were generated by cotransfection of 293T cells with pNL4-3.Luc.R-E-backbone and viral envelope protein expression plasmids as previously described^18^. Mouse anti-human CD32 monoclonal antibody (clone number FLI8.26) was purchased from BD Pharmingen (USA).

### ADE of pseudotyped SARS-CoV-2 infection of Raji cells, K562 cells, and primary B cells

The ADE effect of plasma and antibodies was measured by *in vitro* enhancement of pseudotyped SARS-CoV-2 infection with Raji cells, K562 cells and primary B cells. Briefly, 50 μl of Raji cells or K562 cells were seeded into a 96-well plate pre-coated with 100 μl of 0.1 mg/ml Poly L-lysine at a concentration of 2 × 10^4^ cells per well and cultured at 37 °C for 48 hours. For primary B cells ADE assay, 100 μl of cultured B cells were seeded into wells at a concentration of 1 × 10^4^ cells per well in the presence of irritated 3T3-msCD40L feeder cells, IL-2, and IL-21 and cultured at 37 °C for 48 hours. Ten μl of heat-inactivated plasma were two-fold serially diluted with DMEM with 10% FBS and mixed with 40 μl pseudovirus at 37 °C for 30 minutes. For ADE inhibition assay, different concentrations of RBD or S1 protein from SARS-CoV-2, RBD or S1 protein from SARS-CoV, 20-mer peptides spanning RBD region (20 μg/ml), EK1 peptide (50 μM), or chloroquine (50 μM), or mouse anti-human CD32 monoclonal antibody (5 μg/ml) were incubated with serially diluted patient plasma at 37 °C for 1 hour before mixing with pseudovirus. The mixture was added into cultured cells for infection. After 12 hours, 150 μl of culture medium were added to the cells and incubated for an additional 48 hours. The infection of cells was evaluated by luciferase expression, as determined with a luciferase assay system (Promega) and read on a luminometer (Perkin Elmer). The enhancement of virus infection was expressed as the fold changes of luciferase reading comparing to virus control without addition of plasma or antibodies.

### Purification of IgG from human plasma

Heat-inactivated human plasma samples were 1:6 diluted in PBS and filtered through 0.22 µm filters. The diluted plasma was incubated with protein G beads (Smart-Lifesciences) at 4 °C overnight. The mixture was loaded on filtration column, and IgG-depleted plasma was collected from the flow through. After washing with 150 ml PBS, the beads binding IgG were eluted with 8 ml of 0.1M glycine-HCl buffer (pH 2.7) and neutralized with 200 µl of 2 M Tris-HCl buffer (pH 8.0). The eluted IgG was concentrated using Amicon Ultra centrifugation units (50 kDa, Millipore) after triple washing with 15 ml PBS. Purified IgG was diluted with PBS to the same volume as that of the original plasma samples before evaluation.

### Neutralization assay

Neutralization activity of plasma and antibodies was measured by the inhibition of pseudovirus infection with 293 T/ACE2 cells as previously described^18^. Briefly, 293 T/ACE2 cells were seeded in a 96-well plate at a concentration of 10^4^ cells per well and cultured for 12 hours. Then, ten μl heat-inactivated plasma were five-fold serially diluted with DMEM with 10% FBS and mixed with 40 μl of pseudovirus. The mixture was added to cultured 293 T/ACE2 cells for infection. The culture medium was refreshed after 12 hours and incubated for an additional 48 hours. Assays were developed with a luciferase assay system (Promega), and the relative light units (RLU) were read on a luminometer (Perkin Elmer). The titers of NAbs were calculated as 50% inhibitory dose (ID50), expressed as the highest dilution of plasma which resulted in a 50% reduction of luciferase luminescence compared with virus control.

### Cell-cell fusion assay mediated by ADE patient plasma

Cell-cell fusion assay was conducted as previously described with modification^27^. Briefly, HEK-293T cells expressing the SARS-CoV-2 S protein on the cell membrane were used as effector cells, while Raji cells were used as target cells. HEK293T cells were transfected with plasmid pAAV-IRES-EGFP-SARS-2-S, using transfection reagent VigoFect (Vigorous Biotechnology, China). Raji cells were seeded at a density of 5 × 10^4^ cells per well into the 96-well plates which were precoated with 100 μl of 0.1 mg/ml of Poly L-lysine for 30 min at 37°C. The effector cells were collected 24 hours after transfection and mixed with the serially diluted serum at 37°C for 30 min. The mixture of effector cells and serum was applied onto the Raji cells and cultured for an additional 24 hours. After fixing with 4 % paraformaldehyde, the cells were observed and captured using an inverted fluorescence microscope (Nikon Eclipse Ti-S). The fused cells were counted on five random fields in each well. For inhibition assay, EK1 peptide or chloroquine (TargetMol) was two-fold serially diluted in RPMI 1640 and then mixed with the effector cells and serially diluted ADE patient plasma at 37°C for 30 min. Then, the mixture was applied to the Raji cell as described above.

### ELISA

SARS-CoV-2 RBD, S1, or S2 protein were coated on a MaxiSorp Nunc-immuno 96-well plate (Thermo Scientific, USA) overnight at 4 °C. Wells were blocked with 5% nonfat milk (Biofroxx, Germany) in PBS for 1 hour at room temperature, followed by incubation with 1:400 diluted sera or serially diluted sera in disruption buffer (PBS, 5% FBS, 2% BSA, and 1% Tween-20) for 1 hour at room temperature. A 1:2500 dilution of horseradish peroxidase (HRP)-conjugated goat anti-human IgG antibody (Jackson Immuno Research Laboratories, USA) was added for 1 hour at room temperature. Wells were developed using ABST (Thermo Scientific, USA) for 30 minutes and read at 405 nm on a Multiskan FC plate reader (Thermo Scientific, USA).

### Memory B-cell staining, sorting and antibody cloning

SARS-CoV-2-specific monoclonal antibodies were isolated from mononuclear cells (PBMC) of recovered patients by *in vitro* single B cell as previously described^35^. Briefly, CD19^+^IgA^−^IgD^−^IgM^−^ memory B cells were sorted and resuspended in medium with IL-2, IL-21, and irradiated 3T3-msCD40L feeder cells, followed by seeding into 384-well plates at a density of 4 cells per well. After 13 days of incubation, supernatants from each well were screened for neutralization activity using a high-throughput micro-neutralization assay against SARS-CoV-2. From the wells that scored positive in the neutralization assay, the variable region of the heavy chain and the light chain of the immunoglobulin gene was amplified by RT–PCR and re-expressed as described previously^36,37^. The full-length IgG was purified using a protein G column (Smart-Lifesciences).

### Biolayer interferometry binding assay

The kinetics of monoclonal antibody binding to SARS-CoV-2 RBD protein was measured by biolayer interferometry binding assay on a FortéBio OctetRED96 instrument using anti-human IgG (AHC) biosensors as previously described^38^ The assay followed sequential steps at 30°C as follows. First, the biosensor was immersed in sterile water for 60s, and 10 μg/ml of antibody was loaded on the biosensors. The biosensors were dipped into 0.02% PBST (PBS with 0.02% Tween) for 300 s to reach baseline and then incubated with serially diluted RBD protein solutions for association and PBST for dissociation. Results were analyzed, and K_on_, K_off_ and KD were calculated by FortéBio Data Analysis software (Version 8.1) using 1:1 binding and a global fitting model.

### Cryo-EM sample preparation

The peak fractions of complex were concentrated to about 1.5 mg/mL and applied to the grids. Aliquots (3.3 μL) of the protein complex were placed on glow-discharged holey carbon grids (Quantifoil Au R1.2/1.3). The grids were blotted for 2.5 s or 3.0 s and flash-frozen in liquid ethane cooled by liquid nitrogen with Vitrobot (Mark IV, Thermo Scientific). The cryo-EM samples were transferred to a Titan Krios operating at 300 kV equipped with Cs corrector, Gatan K3 Summit detector and GIF Quantum energy filter. Movie stacks were automatically collected using AutoEMation ^39^, with a slit width of 20 eV on the energy filter and a defocus range from −1.2 µm to −2.2 µm in super-resolution mode at a nominal magnification of 81,000×. Each stack was exposed for 2.56 s with an exposure time of 0.08 s per frame, resulting in a total of 32 frames per stack. The total dose rate was approximately 50 e^-^/Å^2^ for each stack. The stacks were motion corrected with MotionCor2 ^40^ and binned 2-fold, resulting in a pixel size of 1.087 Å/pixel. Meanwhile, dose weighting was performed ^41^. The defocus values were estimated with Gctf ^42^.

### Data processing

Particles for all samples were automatically picked using Relion 3.0.6^43-46^ from manually selected micrographs. After 2D classification with Relion, good particles were selected and subjected to two cycles of heterogeneous refinement without symmetry using cryoSPARC ^47^.The good particles were selected and subjected to Non-uniform Refinement (beta) with C1 symmetry, resulting in 3D reconstruction for the whole structures, which were further subjected to 3D auto-refinement and post-processing with Relion. For interface between SARS-CoV-2 S protein and mAb, the dataset was subjected to focused refinement with adapted mask on each RBD-mAb sub-complex to improve map quality. Then the datasets of three similar RBD-mAb sub-complexes were combined and subjected to focused refinement with Relion. The combined dataset was recentered on the interface between RBD and mAb and re-extracted. The re-extracted dataset was 3D classified with Relion focused on RBD-mAb sub-complex. Then the good particles were selected and subjected to focused refinement with Relion, resulting in 3D reconstruction of better quality on the RBD-mAb sub-complex.

The resolution was estimated with the gold-standard Fourier shell correlation 0.143 criterion ^48^ with high-resolution noise substitution ^49^. Refer to Supplemental Figures S6-S7 and Supplemental Table S1 for details of data collection and processing.

### Model building and structure refinement

For model building of all complexes of S-ECD of SARS-CoV-2 with mAb, atomic models (PDB ID: 7C2L) were used as templates, which were molecular dynamics flexible fitted ^50^ into the whole cryo-EM map of the complex and the focused-refined cryo-EM map of the RBD-mAb sub-complex, respectively. The fitted atomic models were further manually adjusted with Coot ^51^. Each residue was manually checked with the chemical properties taken into consideration during model building. Several segments were not modeled because the corresponding densities were invisible. Structural refinement was performed in Phenix^52^ with secondary structure and geometry restraints to prevent overfitting. To monitor the potential overfitting, the model was refined against one of the two independent half maps from the gold standard 3D refinement approach. Then, the refined model was tested against the other maps. Statistics associated with data collection, 3D reconstruction, and model building were summarized in Table S1.

### Statistical analysis

Statistical analyses were carried out using GraphPad Prism 7.0. Data are indicated as median [IQR]. Differences between nominal data were tested for statistical significance by use of Nonparametric paired or unpaired *t* test. Kruskal-Wallis test was used to compare the differences between three or more groups, and Dunn’s multiple comparisons test was used to correct for multiple comparisons. All tests were two-tailed, and P < .05 was considered statistically significant.

### Role of the funding source

The funders of the study had no role in study design, data collection, data analysis, data interpretation, or writing of the report. The corresponding authors had full access to all data in the study and had final responsibility for the decision to submit for publication.

## Data Availability

Correspondence and requests for all data should be addressed to Jinghe Huang

## Declaration of interests

Patents about the monoclonal antibodies 7F3 and 4L12 in this study are pending.

## Contributions

JH, FW, QZ, LL conceived and designed the experiments. ZS, YZ, and TZ collected the samples and clinical information of patients. JH, ML and FW performed ADE experiments, blocking experiments, peptide array, neutralization assay, ELISA, memory B-cell staining, sorting, and antibody cloning. RY, QZ, ZY and LY performed the structural studies. ZL and LL performed cell-cell fusion assay and blocking assay. YDW, TS, XL, ZL, CL, and TY constructed and expressed SARS-CoV-2 pseudovirus mutants and RBD-Fc protein mutants. YDW, YLW, and JH performed biolayer interferometry binding assay. YDW and YM performed ELISA. DL and YM contributed to ADE experiment. KW, DL, and QW expressed SARS-CoV-2 pseudovirus and their mutants and purification antibodies. PJ contributed to B cell sorting. This project was supervised by YZ, TZ, JS, and YMW. JH, FW, QZ, LL, RY, ML, ZL, and YDW analyzed the data and wrote the manuscript.

## Acknowledgments

We thank Prof. Zhengli Shi for helpful discussion and thank Dr. Vanessa M. Hirsch in NIH, USA for reviewing this manuscript. This work was supported by the National Natural Science Foundation of China (31771008 to JH, 31930001 to YZ and FW, 82041025 to SJ, and 31971123 to QZ), the National Major Science and Technology Projects of China (2017ZX10202102 to JH and 2018ZX10301403 to FW and LL), Hundred Talent Program of Shanghai Municipal Health Commission (2018BR08 to JH), Chinese Academy of Medical Sciences (2019PT350002 to JH), Program of Shanghai Academic/Technology Research Leader (20XD1420300 to LL), the Key R&D Program of Zhejiang Province (2020C04001), the SARS-CoV-2 Emergency Project of the Science and Technology Department of Zhejiang Province (2020C03129), the Leading Innovative and Entrepreneur Team Introduction Program of Hangzhou, and the Special Research Program of Novel Coronavirus Pneumonia of Westlake University and Tencent foundation to QZ. We thank all healthcare personnel and staff in the BSL3 lab involved in the collection of patients’ samples at the Shanghai Public Health Clinical Center, the Cryo-EM Facility and Supercomputer Center of Westlake University for providing cryo-EM and computation support and members of the Core Facility of Microbiology and Parasitology (SHMC) of Fudan University, especially Qian Wang for technical support.

## Supplementary Table and Figures

**Table S1.** Cryo-EM data collection and refinement statistics.

|  |  |  |  |  |
| --- | --- | --- | --- | --- |
| Data collection |  |  |  |  |
| EM equipment | Titan Krios (Thermo Fisher Scientific) |  |  |  |
| Voltage (kV) | 300 |  |  |  |
| Detector | Gatan K3 Summit |  |  |  |
| Energy filter | Gatan GIF Quantum, 20 eV slit |  |  |  |
| Pixel size (Å) | 1.087 |  |  |  |
| Electron dose (e-/Å2) | 50 |  |  |  |
| Defocus range (µm) | -1.2 ~ -2.2 |  |  |  |
| Sample | S-ECD+Ab7F3 |  | S-ECD+Ab4L12 |  |
| Number of collected micrographs | 1,877 |  | 1,219 |  |
| Number of selected micrographs | 1,840 |  | 1,199 |  |
| 3D Reconstruction |  |  |  |  |
| Software | Relion 3.0 |  |  |  |
| Sample | Overall | subcomplex | Overall | subcomplex |
| Number of used particles (Overall) | 91,303 | 29,903 | 314,098 | 557,717 |
| Resolution (Å) | 3.3 | 6.3 | 3.0 | 3.5 |
| Symmetry | C1 |  |  |  |
| Map sharpening B-factor (Å2) | -90 |  |  |  |
| Refinement |  |  |  |  |
| Software | Phenix |  |  |  |
| Cell dimensions |  |  |  |  |
| a=b=c (Å) | 313.056 |  |  |  |
| α=β=γ (°) | 90 |  |  |  |
| Model composition |  |  |  |  |
| Protein residues | 3,508 |  | 4,308 |  |
| Side chains assigned | 3,458 |  | 4,308 |  |
| Sugar | 71 |  | 71 |  |
| RMS deviations |  |  |  |  |
| Bond lengths (Å) | 0.009 |  | 0.007 |  |
| Bond Angles (°) | 0.795 |  | 0.720 |  |
| Ramachandran plot statistics (%) |  |  |  |  |
| Preferred | 88.80 |  | 91.26 |  |
| Allowed | 10.90 |  | 8.60 |  |
| Outlier | 0.30 |  | 0.13 |  |

**Figure S1.**
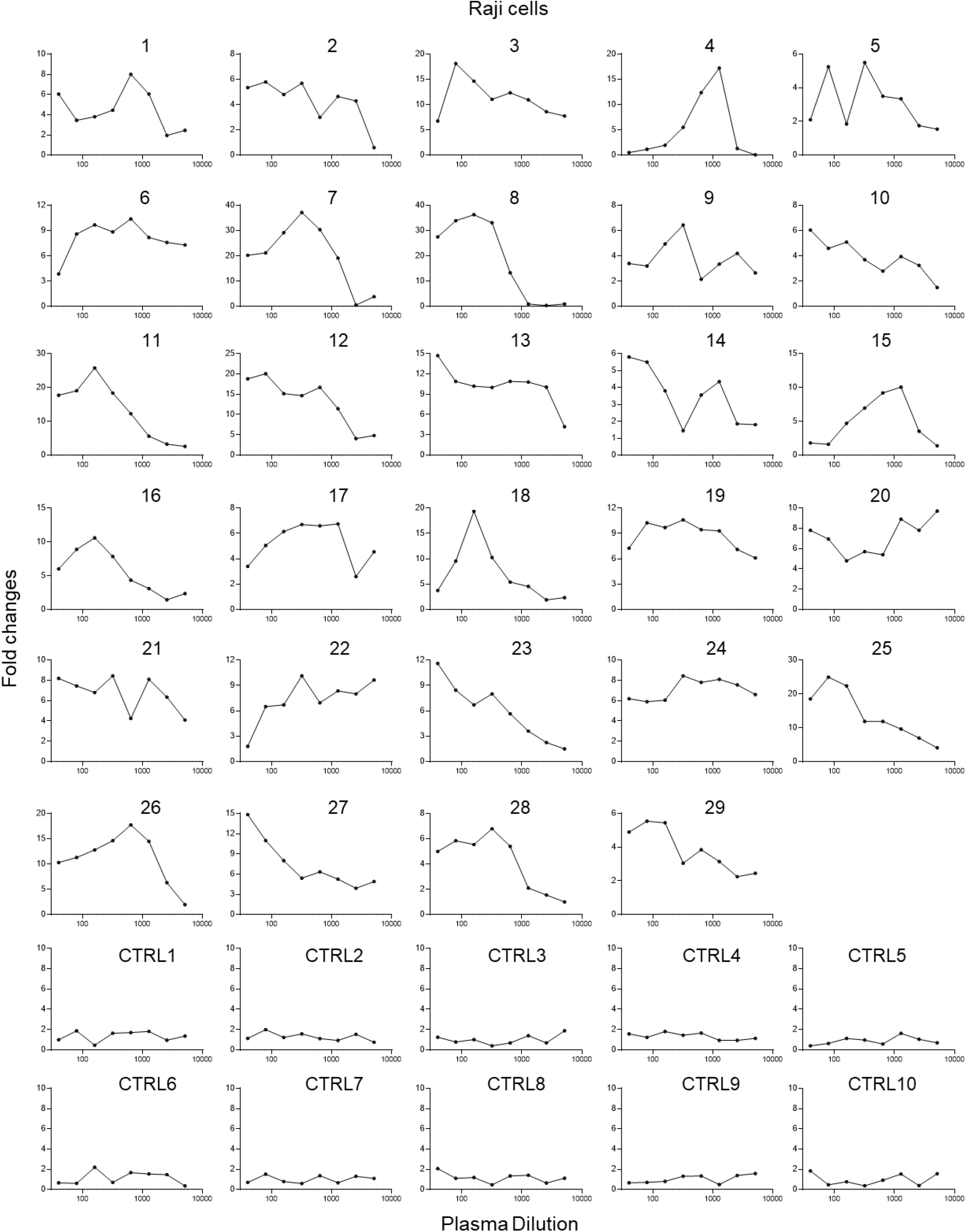
Enhancement of SARS-CoV-2 infection of Raji cells by patient plasma. **(A)** 29 plasma from patients recovered from mild (1-16) and severe COVID-19 (17-29) showed enhancement of SARS-CoV-2 infection of Raji cells. Ten plasma from uninfected donors were used as controls. Plasma samples were two-fold serially diluted and tested for ADE of SARS-CoV-2 infection of Raji cells. The assay was performed in duplicate wells and mean fold changes of luciferase reading comparing to virus control are shown.

**Figure S2.**
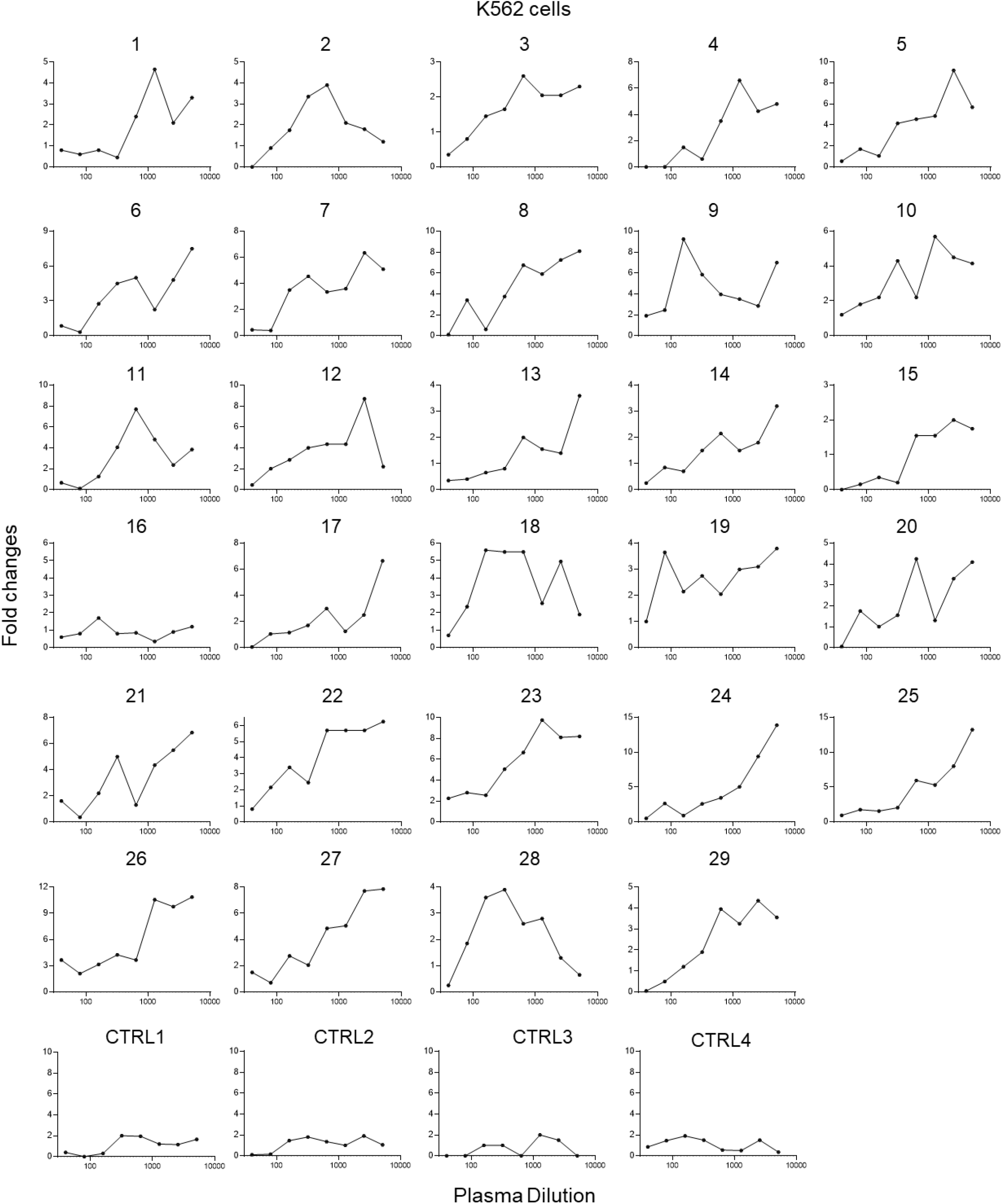
Enhancement of SARS-CoV-2 infection of K562 cells by COVID-19 patient plasma. 29 plasma from patients who recovered from mild (1-16) and severe COVID-19 (17-29) showed enhancement of SARS-CoV-2 infection of K562 cells. Four plasma from uninfected donors were used as controls. The assay was performed in duplicate wells and mean fold changes of luciferase reading comparing to virus control are shown.

**Figure S3.**
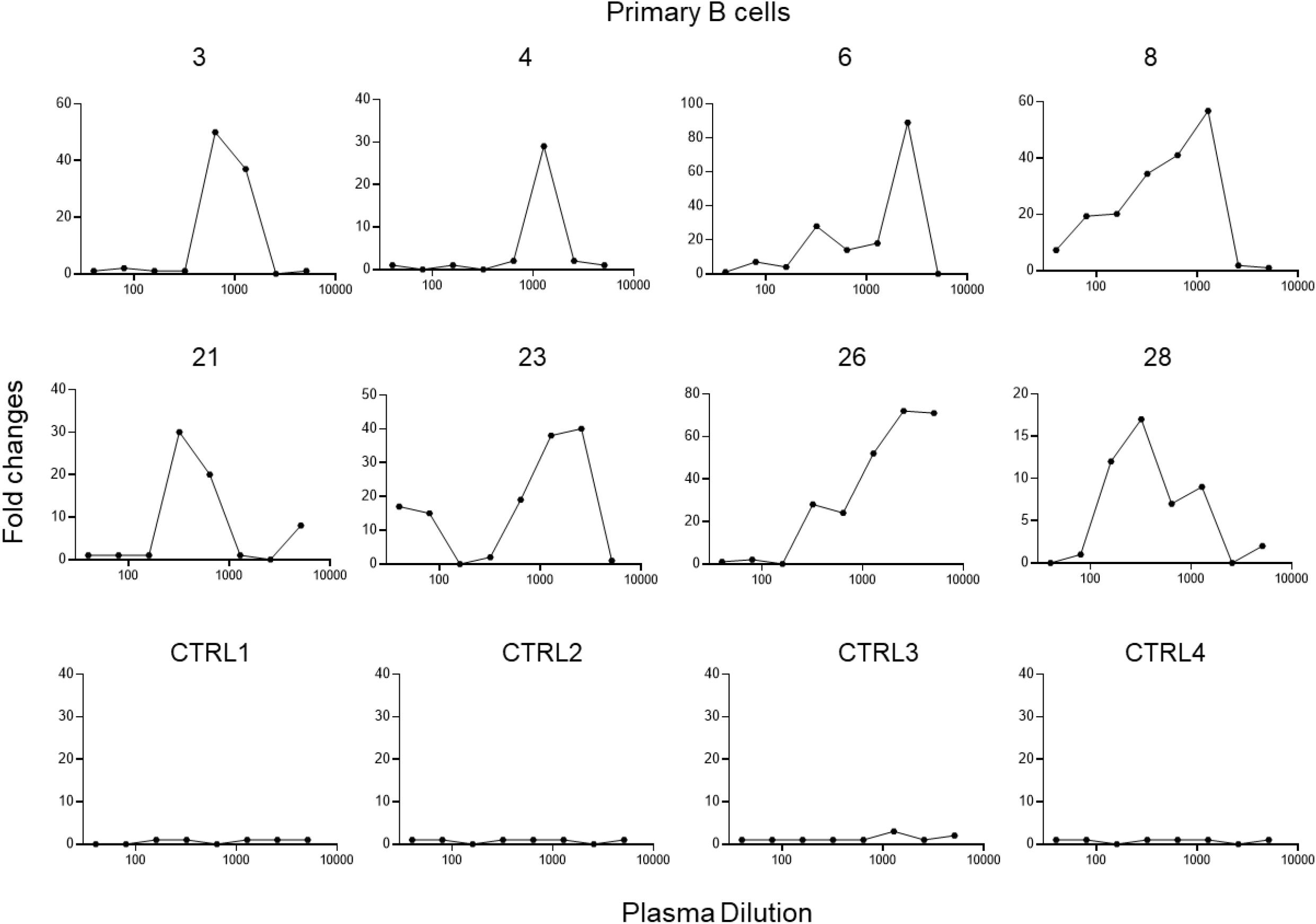
Enhancement of SARS-CoV-2 infection of primary B cells by COVID-19 patient plasma. Eight representative plasma from patients with mild COVID-19 (3, 4, 6, and 8) and severe COVID-19 (21, 23, 26, and 28) showed enhancement of SARS-CoV-2 infection of primary B cells. Four plasma from uninfected donors were used as controls.

**Figure S4.**
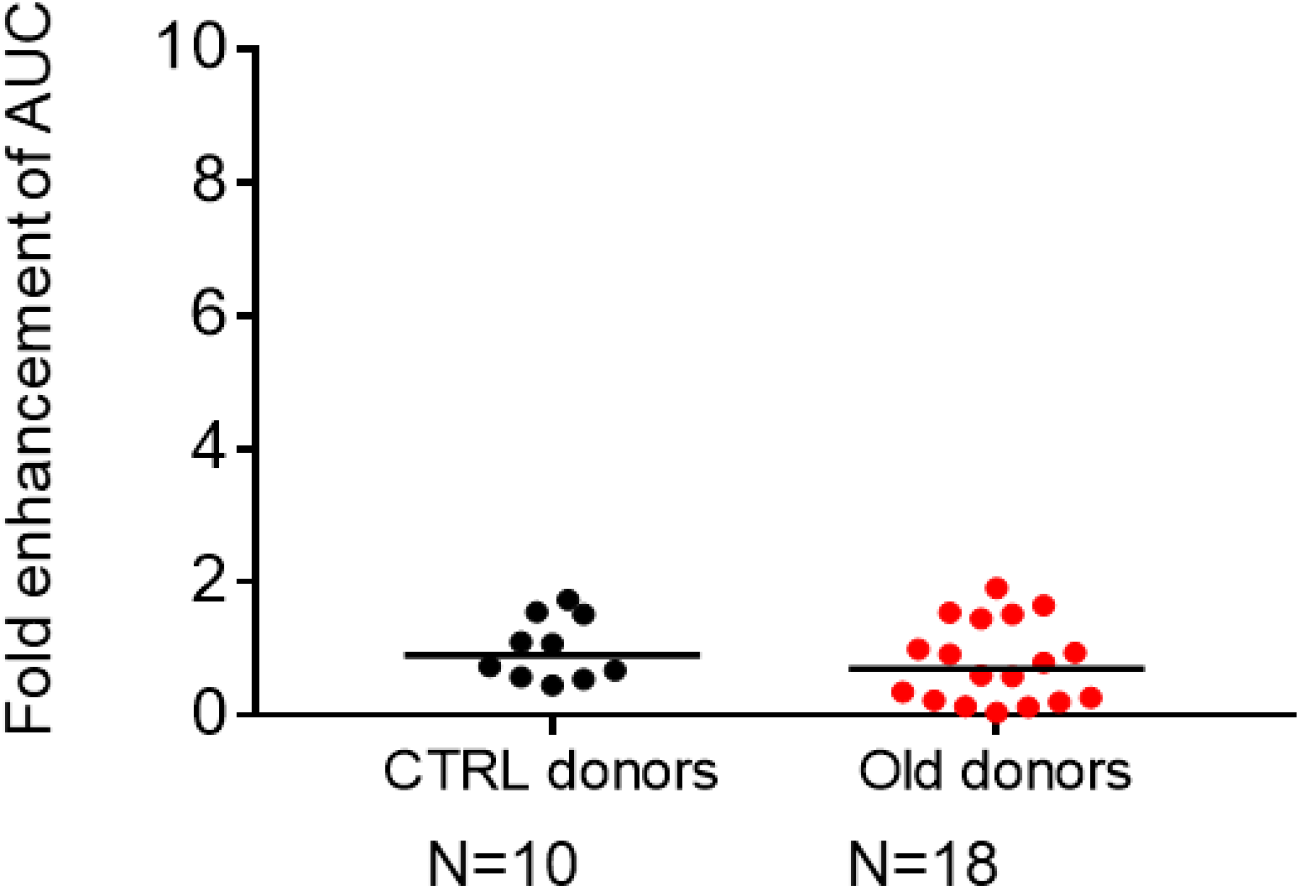
Comparison of ADE effect in plasma from uninfected control donors and uninfected older donors.

**Figure S5.**
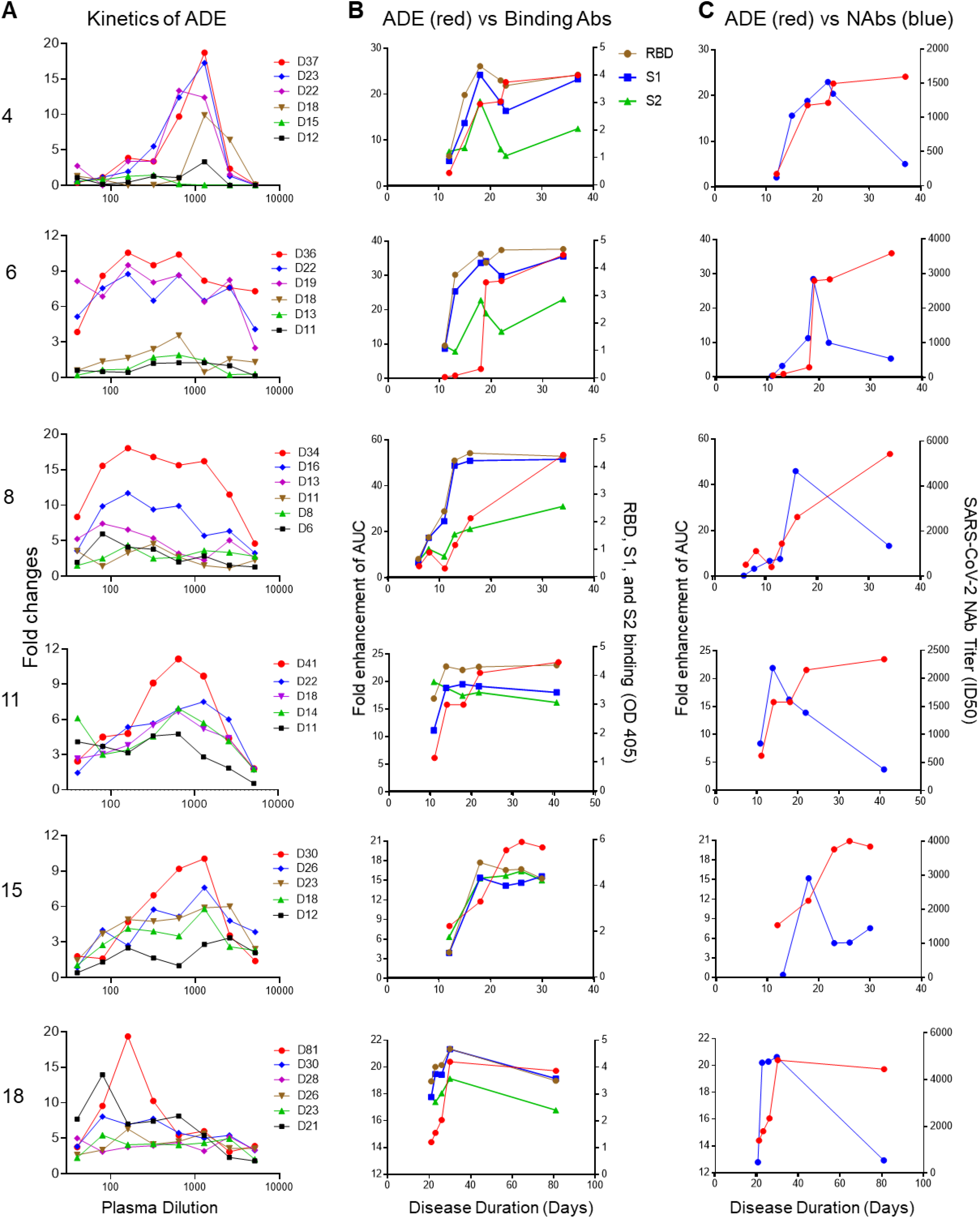
Kinetics of SARS-CoV-2 ADE, spike-binding antibodies and NAbs. **(A)** Kinetics of SARS-CoV-2 ADE effect of six ADE patients are shown. Kinetics of spike-binding antibodies (right Y axis), targeting RBD (brown), S1 (green), and S2 (blue) **(B)** and kinetics of NAbs (right Y axis, blue) **(C)** in six COVID-19 patient plasma are shown and compared with the kinetics of ADE effect (left Y axis, red) in the same patient. 1:400 diluted plasma was incubate with RBD, S1, or S2 protein.

**Figure S6.**
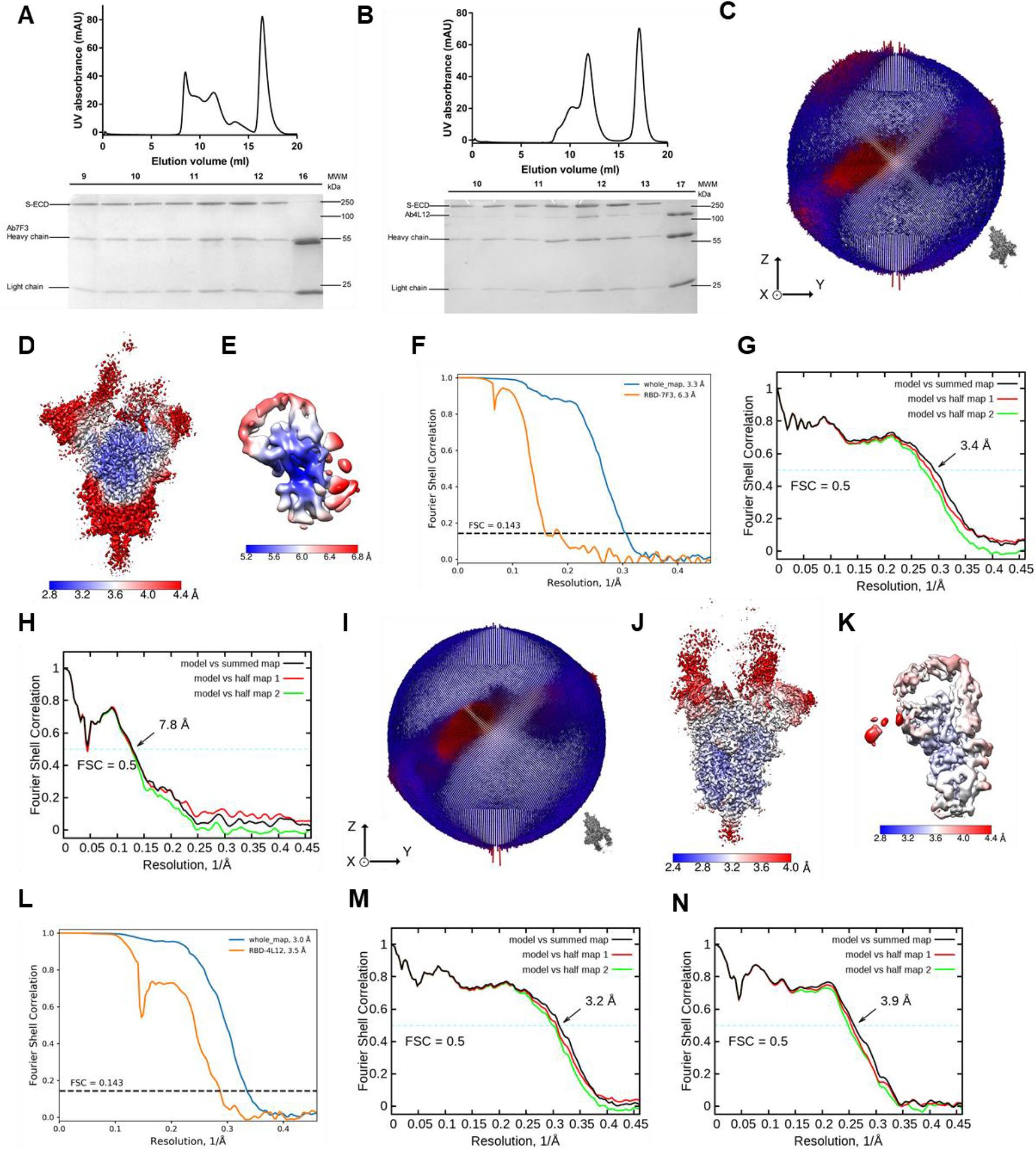
Cryo-EM analysis of S-ECD of SARS-CoV-2 bound with 4A8 complex. **(A)-(B)** Representative SEC purification profile of the S-ECD of SARS-CoV-2 in complex with Ab7F3 and Ab4L12, respectively. **(C)** Euler angle distribution in the final 3D reconstruction of S-ECD of SARS-CoV-2 bound with Ab7F3 complex. **(D)** and **(E)** Local resolution maps for the 3D reconstruction of the overall structure and RBD-Ab7F3 subcomplex, respectively. **(F)** FSC curve of the overall structure (blue) and RBD-Ab7F3 subcomplex (orange). **(G)** FSC curve of the refined model of S-ECD of SARS-CoV-2 bound with Ab7F3 complex versus the overall structure against which it is refined (black); of the model refined against the first half-map versus the same map (red); and of the model refined against the first half-map versus the second half-map (green). The small difference between the red and green curves indicates that the refinement of the atomic coordinates lacks sufficient overfitting. **(H)** FSC curve of the refined model of RBD-Ab7F3 subcomplex, which is the same as **(G). (I)-(N)** is the same as **(C)-(H)**, except for SARS-CoV-2 bound with Ab4L12 complex.

**Figure S7.**
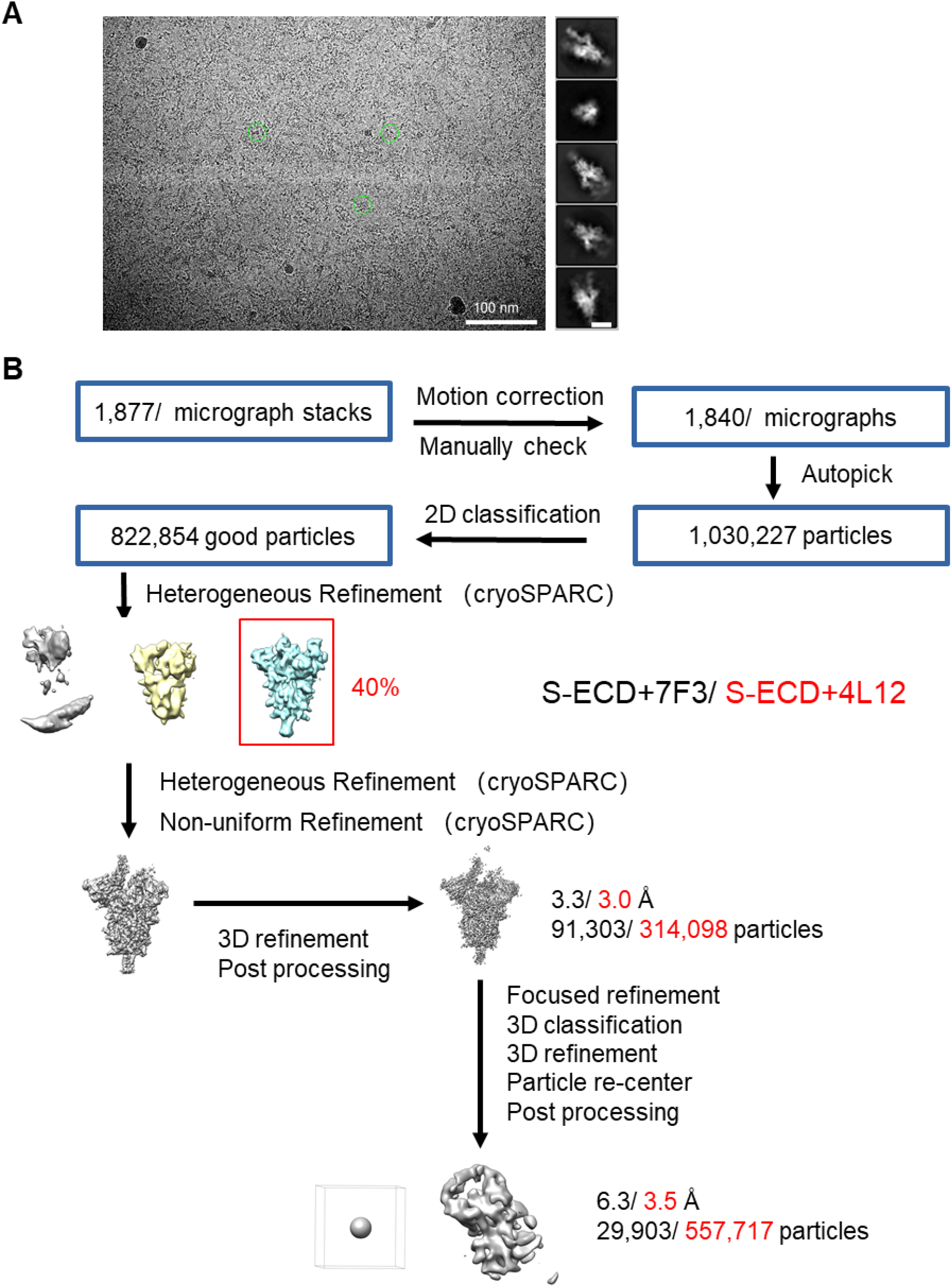
Representative cryo-EM image and flowchart for cryo-EM data processing. **(A)** Representative cryo-EM micrograph and 2D class averages of cryo-EM particle images. The scale bar in 2D class averages is 10 nm. **(B)** Please refer to the ‘Data Processing’ section in Methods for details.

**Figure S8.**
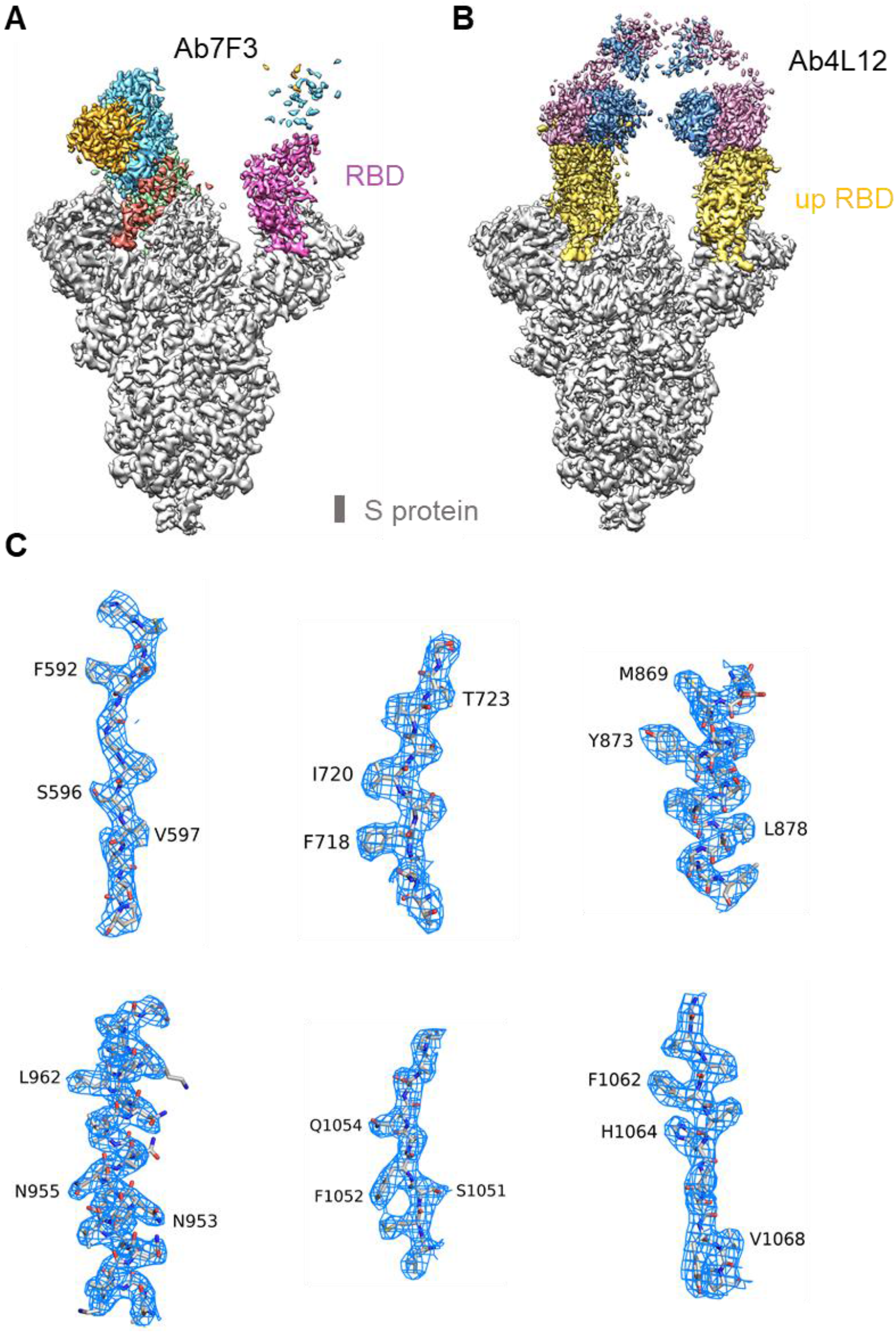
Overall maps of S-ECD in complex with mAb and representative cryo-EM density maps. **(A)** The domain-colored cryo-EM maps of S-ECD of SARS-CoV-2 bound with Ab7F3 complex. **(B)** The domain-colored cryo-EM maps of S-ECD of SARS-CoV-2 bound with Ab4L12 complex. **(C)** Cryo-EM density map for S-ECD in complex with Ab7F3 is shown at threshold of 6 σ.

**Figure S9.**
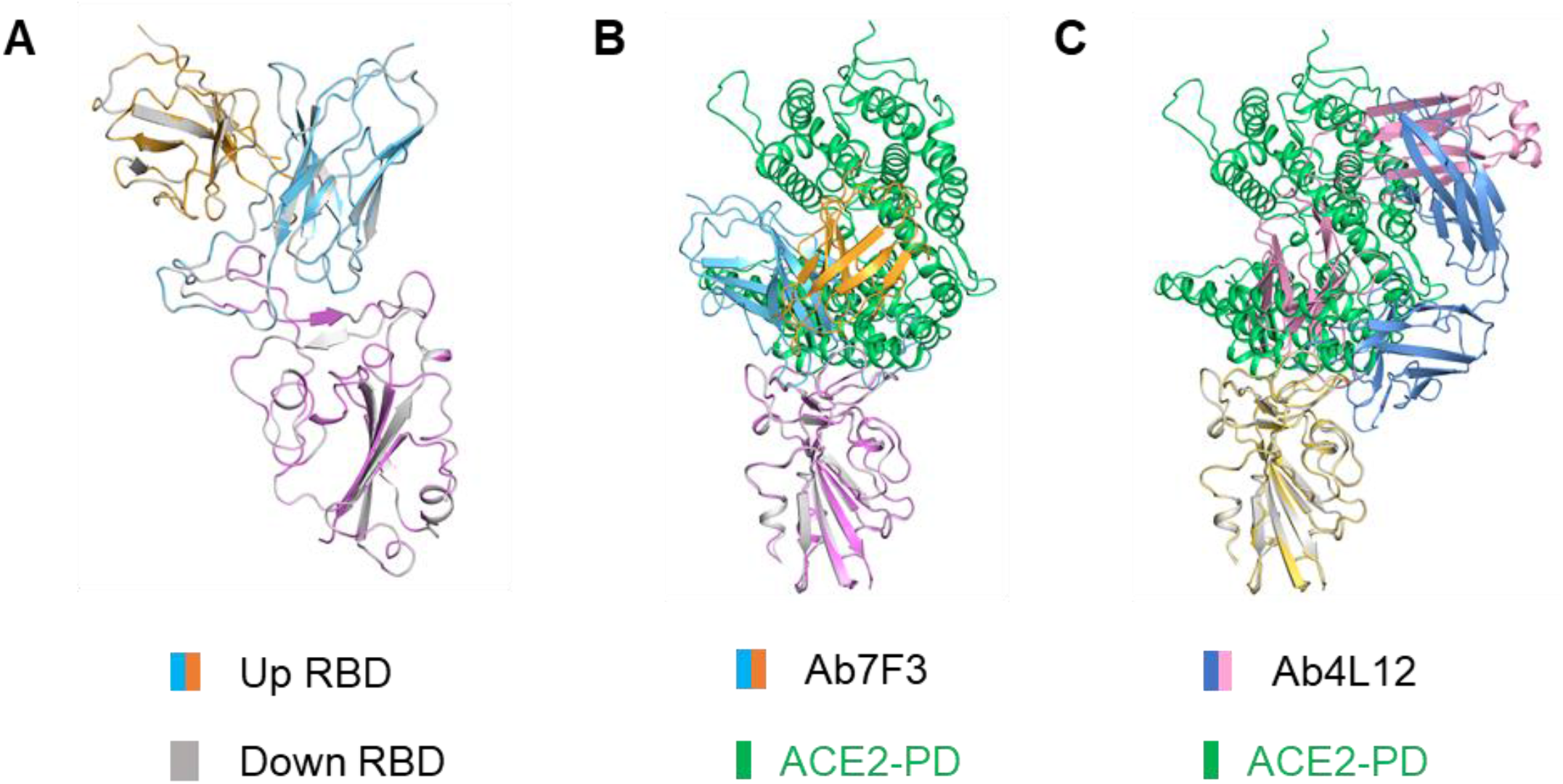
Structural alignment between RBD-mAb subcomplex and RBD-PD complex (PDB ID: 6M0J). **(A)** Superposition in local structure of “up” RBD-Ab7F3 subcomplex and “down” RBD-Ab7F3 subcomplex, indicating no difference between the two maps. **(B)** Structural alignment between RBD-Ab7F3 subcomplex and RBD-PD complex (PDB ID: 6M0J). **(C)** Structural alignment between RBD-Ab7F3 subcomplex and RBD-PD complex (PDB ID: 6M0J).

**Figure S10.**
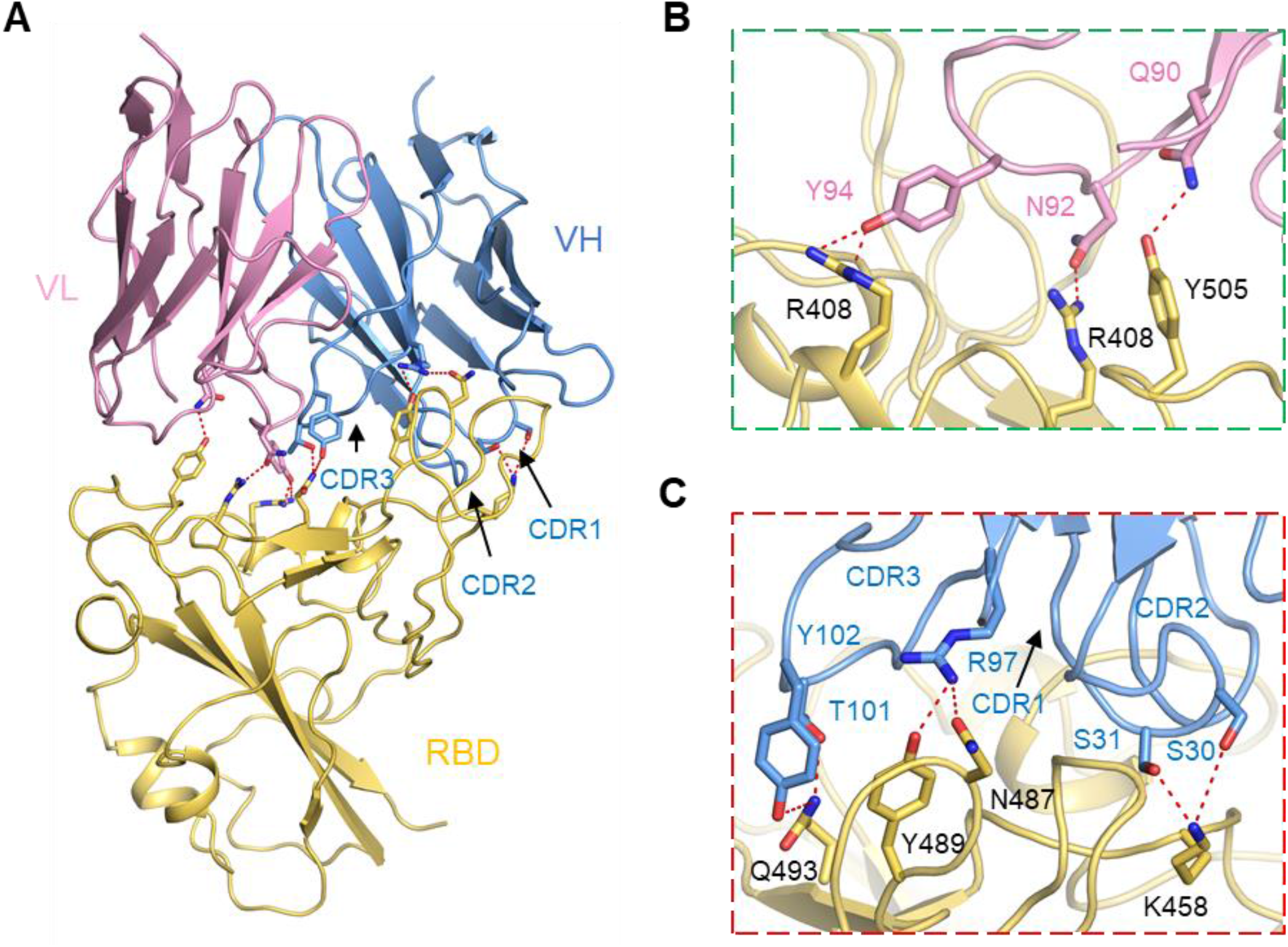
Interactions between the RBD and Ab4L12. **(A)** Extensive hydrophilic interactions on the interface between RBD and Ab4L12. **(B)** and **(C)** Detailed analysis of the interface between RBD and Ab 4L12. Polar interactions are indicated by red dashed lines.

## Notes

### Author Declarations

The study was conducted under a clinical protocol approved by the Investigational Review Board in the Shanghai Public Health Clinical Center (Study number: YJ-2020-S018-02).

